# A Benchmark of methods for SARS-CoV-2 whole genome sequencing and development of a more sensitive method

**DOI:** 10.1101/2024.10.09.24313595

**Authors:** Anthony Bayega, Sarah J. Reiling, Isabelle Dubuc, Annie Gravel, Louis Flamand, Jiannis Ragoussis

## Abstract

The raging COVID-19 pandemic caused by SARS-CoV-2 has so far claimed the lives of 4.6 million people and continues to infect many more. Further, virus evolution has caused mutations that have compromised public health interventions like vaccination regimes and monoclonal antibody and convalescent sera treatments. In response, unprecedented large-scale whole genome viral surveillance approaches have been devised to keep track of the evolution and transmission patterns of the virus within and across populations. Here, we aimed to compare efficiencies of SARS-CoV-2 whole genome sequencing approaches using synthetic SARS-CoV-2 genome and six cell culture SARS-CoV-2 variants titrated to represent samples at high, medium, and low viral load. We found that the ARTIC protocols performed best in terms of PCR amplicon yield returning 67% more amplicons than Entebbe protocol which was the second highest PCR amplicon yielding protocol. ARTIC v4.1 protocol yields were only slightly better than ARTIC v3. Despite yielding the lowest PCR amplicons, the SNAP protocol showed the highest genome completeness using a synthetic genome at high viral titre followed by ARTIC protocols. However, the ARTIC protocols showed highest genome completeness with cell culture SARS-CoV-2 variants across high, medium and low viral titres. ARTIC protocol also performed best in calling the correct lineage among cell culture SARS-CoV-2 variants across different viral titres. We also designed a new method termed ARTIC-Amp which leverages ARTIC protocol and performs a rolling circle amplification to increase yield of amplicons. In a proof-of-principle experiment, this method showed 100% coverage in all four targeted genes across three replicates unlike the ARTIC protocol missed one gene in two of the three replicates. Our results demonstrate the robustness of the ARTIC protocol and propose an improved method that could be useful for samples that routinely have limited SARS-CoV-2 RNA such as wastewater samples.

**Contribution to the field**

## Introduction

As of September 2022, the current coronavirus disease 2019 (COVID-19) pandemic has resulted in at least 603 million infections and 6.48 million deaths worldwide since its inception in December 2019 (WHO). The causative agent of COVID-19 was quickly identified as a respiratory virus of Betacorona genus and named severe acute respiratory syndrome corona virus 2 (SARS-CoV-2) (Wu et al., 2020; Zhu et al., 2020). This enveloped virus is composed of a 29903 basepair positive single stranded RNA genome that encodes four structural (Spike, Membrane, Envelope, and Nucleocapsid) and at least 29 non-structural proteins (Lu et al., 2020; Wu et al., 2020). The NSP12 gene encodes an RNA-dependent RNA polymerase (RdRp) which is responsible, along with other viral and host co-factors, for replication of the viral genome once inside the cell. Despite the proofreading activity of the RdRp conferred by NSP14, the evolutionary rate of SARS-CoV-2 measured as inter-host temporal variation in consensus sequence is estimated at 1.5±0.5 x 10^-3^ per site per year (Domingo et al., 2021). This viral evolution creates mutations that are of huge public health concern. Indeed, it was noted in November 2020 that new mutations were arising that led to more transmissible and vaccine evading variants of SARS-CoV-2 (Davies et al., 2021; Thorne et al., 2022; Walker et al., 2021). This alerted scientists of the need to perform large-scale whole genome sequencing (WGS) as a surveillance method. Large scale WGS in turn elucidated the worldwide spread of variants and led to identification of new variants. Currently SARS-CoV-2 variants are of three classes: variants of concern (VOCs), variants of interest (VOIs), and variants under monitoring (VUMs). VOCs include the Alpha (B1.1.7), Beta (B1.351), Gamma (P1), Delta (B1.617.2), and Omicron (BA.1) (Harvey et al., 2021; Mendiola-Pastrana et al., 2022). Viral surveillance is critical to track the evolution and transmission patterns of these VOC within and across communities and quickly identify new mutations and variants. Further, surveillance facilitates tracking of the impact of mutations on public health and public health interventions such as vaccinations, monoclonal and convalescent sera treatment, drugs, and personal hygiene and protection interventions like masks and number of metres required for social distancing.

Real-time quantitative PCR (RT-qPCR) is the gold standard surveillance method with very high sensitivity. This quantitative method provides a cycle threshold metric (Ct value) that provides a linear inverse relationship with amount of viral RNA in a sample. The drawback of RT-qPCR, however, is that no sequence information is produced. Whole genome sequencing methods have thus been used. A widely used SARS-CoV-2 WGS protocol is one developed by an international workgroup comprising scientist from UK, Belgium, and USA called Advancing Real-Time Infection Control network (ARTIC). Primers used in the ARTIC protocol have undergone at least three major revisions in response to SARS-CoV-2 evolution giving rise to versions 3, 4, and 4.1. Several other SARS-CoV-2 whole genome sequencing protocols have been made available either by academic research labs or companies. Although benchmarking studies have been attempted (Liu et al., 2021; Plitnick et al., 2021), it is not clear how the different methods compare to each other and how they perform with low viral titre samples such as environmental samples.

Despite their low viral titre, environmental samples have shown promise as an alternative to clinical samples for early detection of SARS-CoV-2 variants. Municipal wastewater surveillance, for example, has potential to provide passive population-scale surveillance and has been shown to identify new SARS-CoV-2 variants up to three weeks before clinical identification (Ahmed et al., 2021; Larsen & Wigginton, 2020; Smyth et al., 2022). One of the biggest drawbacks of environmental samples, particularly municipal wastewaters, is the very low amounts of SARS-CoV-2 with Ct values routinely above 35 (Jafferali et al., 2021). This can be attributed partly to low recovery rates of methods used to extract nucleic acids that can be as low as 0-25 % and presence of inhibitors of molecular assays in extracted wastewater samples (Jafferali et al., 2021). It is thus imperative to develop very sensitive methods to enable reliable early detection of SARS-CoV-2.

Here, we aimed to compare the performance of five SARS-CoV-2 whole genome sequencing protocols including the ARTIC versions 3 and 4.1, QIAseq DIRECT SARS-CoV-2 (Qiagen, Hilden, German), Swift Normalase Amplicon Panel for SARS-CoV-2 additional genome completeness (SNAP, Swift Bioscience), Midnight protocol (Nikki E. Freed et al., 2020), and Entebbe protocol (Cotten et al., 2020). After finding that all protocols performed poorly with low viral titre samples we sought to design a custom protocol. In the custom protocol which we termed ARTIC-Amp (for amplified ARTIC), we leveraged the ARTIC v4.1 protocol which we found to perform best by using amplicons generated from the ARTIC v4.1 protocol to perform another round of isothermal rolling circle amplification as applied recently (Volden et al., 2018). In a proof-of-principle experiment, the ARTIC-Amp protocol showed improved performance over regular ARTIC protocol.

## Materials and methods

### Sample preparation: synthetic SARS-CoV-2 genome

We purchased the Twist Synthetic SARS-CoV-2 RNA Control 1 (102019, Twist Bioscience, CA, USA). This control consists of six non-overlapping 5 kb ssRNA fragments covering 99.9 % of the viral genome (GenBank ID MT007544.1, GISAID NAME Australia/VIC01/2020) and is reconstituted at 1,000,000 copies per microliter. We serial diluted the control in water to generate solutions at 1×10^6^, 1×10^5^, 1×10^4^, 1×10^3^, and 1×10^2^ copies/mL. These were stored at −80 °C.

### Sample preparation: cell culture SARS-CoV-2 virus

We obtained SARS-CoV-2 virus samples as follows: B.1 and B.1.1.7 were obtained from the Laboratoire de Santé Publique du Québec (LSPQ), B.1.351 and P.1 were obtained from BEI Resources (VA, USA), B.1.617.2 was obtained from Canada’s National Microbiology Laboratory (NML), and BA.1 was obtained from British Columbia Centre for Disease Control (BCCDC, Canada). The samples were cultured in VERO cells at a multiplicity of infection of 0.002 for 4 – 5 days. The supernatants were collected and centrifuged to remove cells and debris. Total RNA was extracted using the Bead Mill Tissue RNA purification kit (26-010B, OMNI International, GA, USA). Briefly (see full protocol in supplementary materials), 300 µL of supernatant were mixed with ceramic beads, 300 µL of RLB buffer, 10 µL of antifoam reagent, and 12 µL of β-mercaptoethanol. Following homogenization, the solution was centrifuged, reconstituted with one volume of 70 % ethanol and vortexed. The solution was then transferred to an Omni RNA column, where the RNA was hybridized to the column, washed, and eluted in DEPC-treated water. RNAs were reverse transcribed to cDNA using SuperScript™ IV VILO™ mastermix, as per manufacturer’s recommendations (ThermoFisher Scientific). Quantitative real-time PCR (RT-qPCR) was then performed to determine cycle threshold values (Ct, Table 1). SARS-CoV-2 Envelope (E) gene forward (5’-ACAGGTACGTTAATAGTTAATAGCGT-3’) and reverse (5’-ATATTGCAGCAGTACGCACACA-3’) primers, along with the SsoAdvanced Universal SYBR Green Supermix (Bio-Rad Laboratories Ltd) were used. RNAs were also diluted at 1:100 and quantified by digital droplet PCR (ddPCR) using the One-Step RT-ddPCR Advanced kit for Probes (Bio-Rad Laboratories Ltd) following manufacturer’s recommendations. Primers used for ddPCR included the SARS-CoV-2 E primers used for qPCR experiments and the SARS-CoV-2 probe (5’-ACACTAGCCATCCTTACTGCGCTTCG-3’). All samples were normalized to 1000 copies/µL and then further log-serial diluted to generate new samples at 100, 10, 1, and 0.1 copies/µL. Eleven microliters of each sample was then used as input template for reverse transcription. For the main experiments, we used samples at 100, 1, and 0.1 copies/µL to simulate clinical samples at high viral load, medium viral load, and low viral load, respectively.

**Table 1:** Cell culture wildtype SARS-CoV-2 and its variants used in this study and their RT-qPCR cycle threshold (Ct) values and digital droplet PCR (ddPCR) concentrations. (ND = not determined)

| No. | WHO label | PANGO lineage | First detection | Ct value | ddPCR Conc'n (copies/μL) |
| --- | --- | --- | --- | --- | --- |
| 1. | Wild-type | B.1 | WU | 15.21 | $2.1 \times 10^5$ |
| 2. | Alpha | B.1.1.7 | UK | 12.06 | $9.5 \times 10^5$ |
| 3. | Beta | B.1.351 | South Africa | 11.19 | $2.8 \times 10^6$ |
| 4. | Gamma | P.1 | Brazil | 10.41 | $5.8 \times 10^6$ |
| 5. | Delta | B.1.617.2 | India | 15.46 | $1.3 \times 10^5$ |
| 6. | Omicron | BA.1 | South Africa | 12.62 | ND |

### ARTIC protocol

We used the ARTIC workflow as described by the authors (https://www.protocols.io/view/ncov-2019-sequencing-protocol-bbmuik6w, (Tyson et al., 2020)). Briefly, 11 µL of sample was reverse transcribed using LunaScript RT SuperMix (NEB, MA, USA) in a 20 µL reaction. We used the Q5 Hot Start High-Fidelity 2X Master Mix (NEB, MA, USA) to prepare two separate pools of PCR mixes corresponding to the two primer pools in the ARTIC workflow. We used both ARTIC version 3 and version 4.1 primers. Five micro litres of cDNA mix was directly added to 20 µL of PCR mix and amplified for 36 cycles. Amplicons were purified using SPRI paramagnetic beads (Beckman Coulter, IN, USA) and quantified using either Qubit fluorometer (Thermo Fischer Scientific) or Quant-iT PicoGreen dsDNA Assay Kits (Thermo Fischer Scientific). Samples were barcoded using Native Barcoding Expansion Kit 96 (EXP-NBD196, Oxford Nanopore Technologies, UK). To multiplex samples, we used 150 ng of purified amplicons or 15 µL of sample for samples with less than 10 ng/µL. The multiplexed library was purified once more followed by sequencing on the PromethION (R9.4.1 flow cells, Oxford Nanopore Technologies, UK).

### Swift Normalase Amplicon Panel (SNAP) protocol

The Swift Normalase Amplicon Panel (SNAP, Swift Bioscience) for SARS-CoV-2 Additional Genome Coverage was used according to manufacturer instructions, except were mentioned otherwise in this manuscript. Briefly, cDNA was generated as described for ARTIC protocol and 10 μL of cDNA added directly into each PCR tube followed by 20 μL of a master mix composed of 2 μL Reagent G1 (primer set), 3 μL Reagent G2, and 15 μL Enzyme G3. We used the Low Viral Load Input Recommendations and thus amplified the cDNA for a total of 28 cycles, except for Omicron (BA.1) samples. Amplicons were purified, quantified, pooled, and sequenced as described for the ARTIC protocol.

### Qiaseq protocol

The QIAseq DIRECT SARS-CoV-2 (Qiagen, Hilden, German) protocol, herein referred to as Qiaseq was used according to manufacturer instructions, except for the cDNA synthesis step which was performed as described for the ARTIC protocols. We followed the guidelines for “Samples with broad/unknown range Ct value” and thus performed 29 PCR cycles. Amplicons were purified, quantified, pooled, and sequenced as described for the ARTIC protocol.

### Midnight protocol

The Midnight protocol was carried out as described by its authors (https://www.protocols.io/view/ncov-2019-sequencing-protocol-rapid-barcoding-1200-bgggjttw (Nikki E. Freed et al., 2020)). cDNA synthesis, amplification, purification, pooling, and sequencing were carried out as described for the ARTIC protocol except that during PCR amplification we used 2.5 μL of cDNA for each of the two 1200 bp primer sets and PCR extension temperature was 65 °C as recommended by the Midnight protocol developers.

### Entebbe protocol

The Entebbe protocol (Cotten et al., 2020) was carried out with some changes to the instructions of the developers. Firstly, cDNA synthesis was carried out as described for the ARTIC protocol that uses random hexamers rather than gene-specific primers as instructed in the Entebbe protocol. cDNA amplification, purification, pooling, and sequencing were carried out as described for the ARTIC protocol except that PCR amplification was carried out following the conditions recommended by Entebbe protocol developers (Cotten et al., 2020).

Noteworthy, we used LunaScript RT SuperMix to generate enough cDNA in one batch from each sample that would be used to evaluate all protocols and thus minimise batch effects. The cDNA was used as input and as specified for each protocol: 5 µL for each of the two pools for ARTIC, Entebbe, and custom protocols, 2.5 µL for each of the two pools of Midnight protocol, and 10 µL for the single pool of SNAP protocol. PCR amplification was done as specified for each protocol: 36 cycles for ARTIC, Midnight, Entebbe, and custom protocol, 29 for Qiaseq protocol, and 28 cycles for SNAP (except for BA.1 where samples were amplified for 36 cycles).

### ARTIC-Amp protocol

We adapted the R2C2 protocol developed by Volden et al. (Volden et al., 2018) to create the custom protocol we termed ARTIC-Amp for amplified ARTIC v4.1 (see Supplementary materials for the full protocol). In a proof-of-principle experiment, four primers among the ARTIC version 4.1 primers were selected as shown in Table 2. For each of the left primers we added the adapter sequence “AATGATACGGCGACCACCGAGATCTACAC” and for each of the right primers we added the adapter “AAGCAGTGGTATCAACGCAGAGT”. We also obtained the Lambda_Splint_F “ACTCTGCGTTGATACCACTGCTTAAAGGGATATTTTCGATCGCTTG” and Lambda_Splint_R “ATCTCGGTGGTCGCCGTATCATTTGAGGCTGATGAGTTCCATATTTG” primers to amplify a 330 bp fragment of the Lambda phage genome (see Supplementary materials for the full protocol). We used the Lambda phage shipped in the SQK-LSK108 kit (ONT, UK) as template. In order to compare the ARTIC v4.1 protocol to our custom protocol we followed the ARTIC protocol for cDNA synthesis, amplification, and sequencing using BA.1 (Omicron) and B.1.617.2 (Delta) cell culture variants at 100, 1, and 0.1 viral particles/µL. For the custom protocol, purified PCR amplicons were taken through the R2C2 protocol prior to sequencing. Briefly, to circularize the amplicons a 12 µL reaction composed of 2x NEBuilder HiFi DNA Assembly Master Mix (NEB, USA) and PCR amplicons and Lambda splint mixed in a molar ratio of 1:3 up to a total of 200 ng of PCR amplicons was incubated at 55 °C for 60 minutes. Non-circularized molecules were removed by exonuclease followed by overnight rolling circle amplification using Phi29 polymerase. T7 Endonuclease was used to de-branch the DNA. The debranched DNA was purified and sequenced as described for the ARTIC protocol.

**Table 2:**
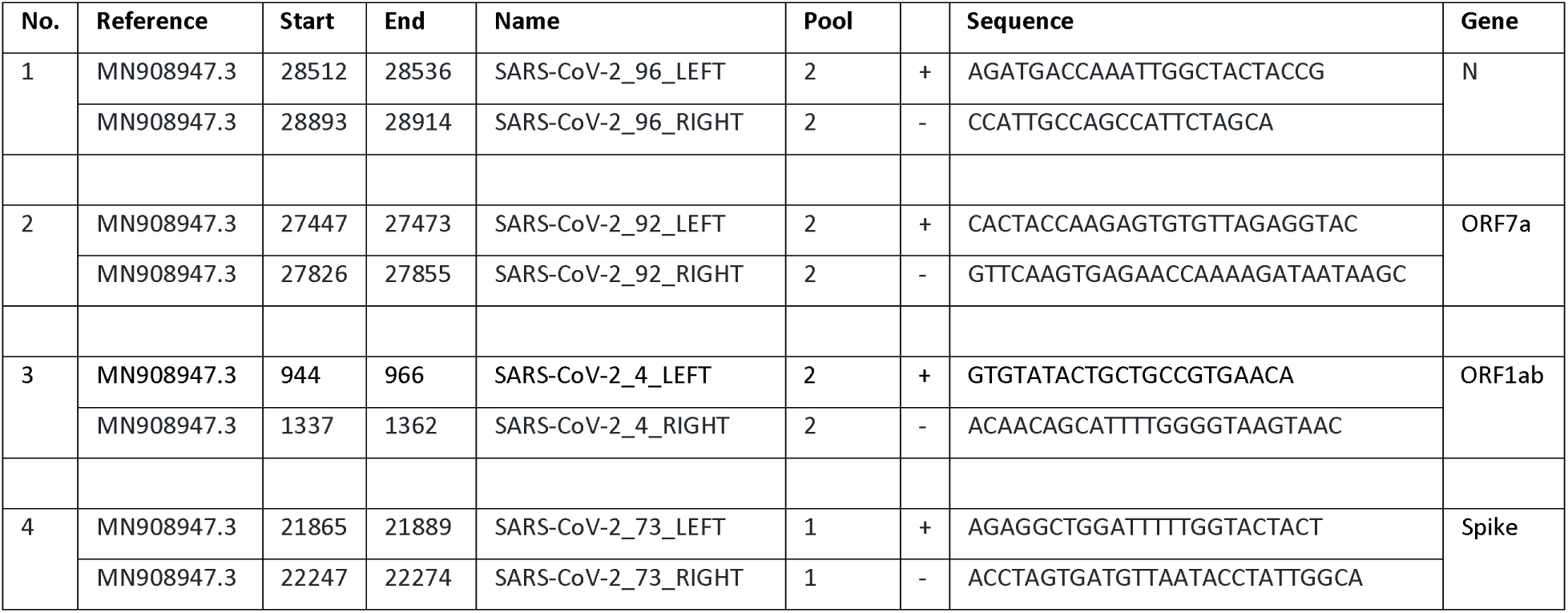
Primers used in the proof-of-principle experiment with our in-house ARTIC-Amp protocol.

## Data analysis

### Basecalling and read processing

All samples were sequenced and basecalled on PromethION with the following parameters; MiniKNOW version: 21.05.20, flow cell: FLO-PRO002, library kit: SQK-LSK109, basecalling type: High accuracy, barcoding kit: EXP-NBD193, Guppy version: 5.0.17. The resulting reads were processed using pychopper (https://github.com/nanoporetech/pychopper) to trim Nanopore sequencing adapters and split chimeric reads containing adapters within them. Processed reads were supplied to the ARTIC pipeline to perform read filtering, primer trimming, amplicon coverage normalisation, variant calling, and consensus building via the Medaka workflow. The specific lineage for each consensus sequence was assigned using the Phylogenetic Assignment of Named Global Outbreak (PANGO) package (Rambaut et al., 2020). To produce and plot statistics from the alignment files, we used the SAMtools (Li et al., 2009) packages ampliconstats and plot-ampliconstats (http://www.htslib.org/doc/samtools-ampliconstats.html), respectively.

## Results

### Short-read protocols show highest genome completeness on synthetic genome

We obtained the Twist Synthetic SARS-CoV-2 RNA Control 1 whose commercial concentration is 1,000,000 particles per microlitre. We performed log serial dilution to obtain samples at 1 x 10^6^, 1 x 10^5^, 1 x 10^4^, 1 x 10^3^, and 1 x 10^2^ particles per millilitre. We processed these samples using five different protocols in triplicate: ARTIC version 3, ARTIC v4, Midnight, SNAP, and Entebbe. ARTIC v3, ARTIC v4, and SNAP protocols are short read protocols yielding amplicons of approximately 450, 450, and 350 bp, respectively while Midnight and Entebbe are long read protocols yielding reads of ∼1200 and 1800 bp, respectively (Supplementary Figure 1). We observed a higher amount of PCR amplicons using the ARTIC V4 protocol compared to all other protocols. For examples, in the highest viral titre samples with 1 x 10^6^ particles/mL we obtained a mean of 162 ng/µL with ARTIC v4 compared to 137 ng/µL, 107 ng/µL, 70 ng/µL, and 8.4 ng/µL with Entebbe, ARTIC v3, Midnight, and SNAP protocols, respectively (Supplementary Figure 2). We then sub-sampled 40,000 reads and used the ARTIC pipeline to reconstruct the genome. We assessed genome completeness measured as the percentage of non ‘N’ bases in the reconstructed genome of the whole genome (Figure 2). The SNAP protocol showed the highest genome completeness at all tested viral particle concentrations. The SNAP protocol showed an average of 96.45 % coverage across the genome. This was followed by ARTIC V3, ARTIC V4, Midnight, and Entebbe. The Midnight protocol was only tested at 1×10^5^ particles/mL.

**Figure 1:**
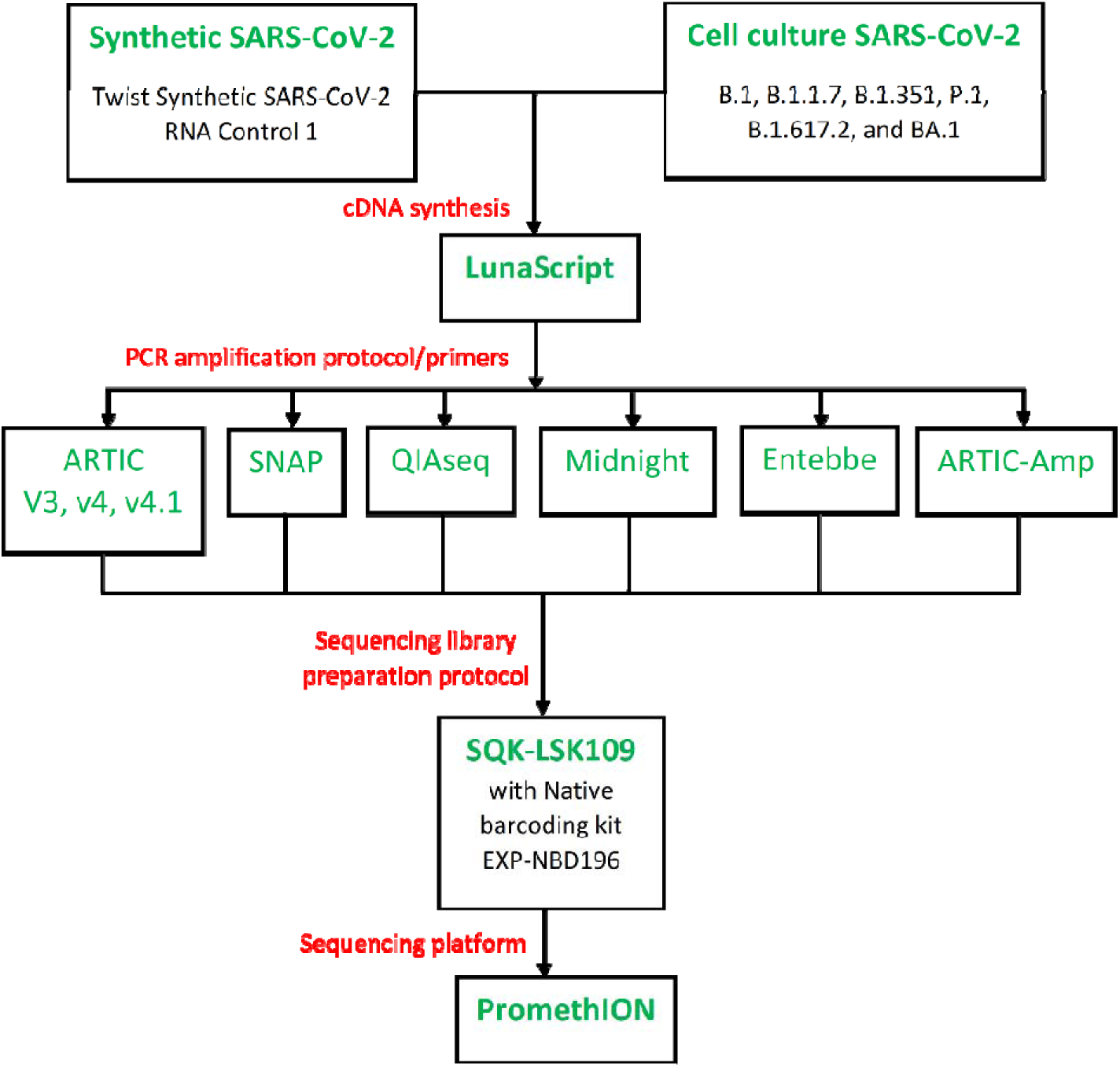
Overview of the study design. Samples (serial diluted synthetic genome or cell culture SARS-CoV-2) were reverse transcribed using LunaScript RT SuperMix Kit (NEB, USA). Ample cDNA per sample was prepared in one batch to test all protocols. cDNA was then processed with a total of six protocols including ARTIC protocol (Tyson et al., 2020), QIAseq DIRECT SARS-CoV-2 (Qiagen, Hilden, German), Swift Normalase Amplicon Panel for SARS-CoV-2 additional genome coverage (SNAP, Swift Bioscience), Midnight protocol (Nikki E. Freed et al., 2020), and Entebbe protocol (Cotten et al., 2020). Our in-house protocol called ARTIC-Amp included a target enrichment step of rolling circle amplification as implemented recently (Volden et al., 2018). cDNA amplification employed protocol-specific primers. For sequencing, we followed the ARTIC protocol except for Qiaseq and SNAP protocols which followed manufacturer directions. Sequencing was performed on the PromethION following Oxford Nanopore Technologies SQK-LSK109 protocol.

**Figure 2:**
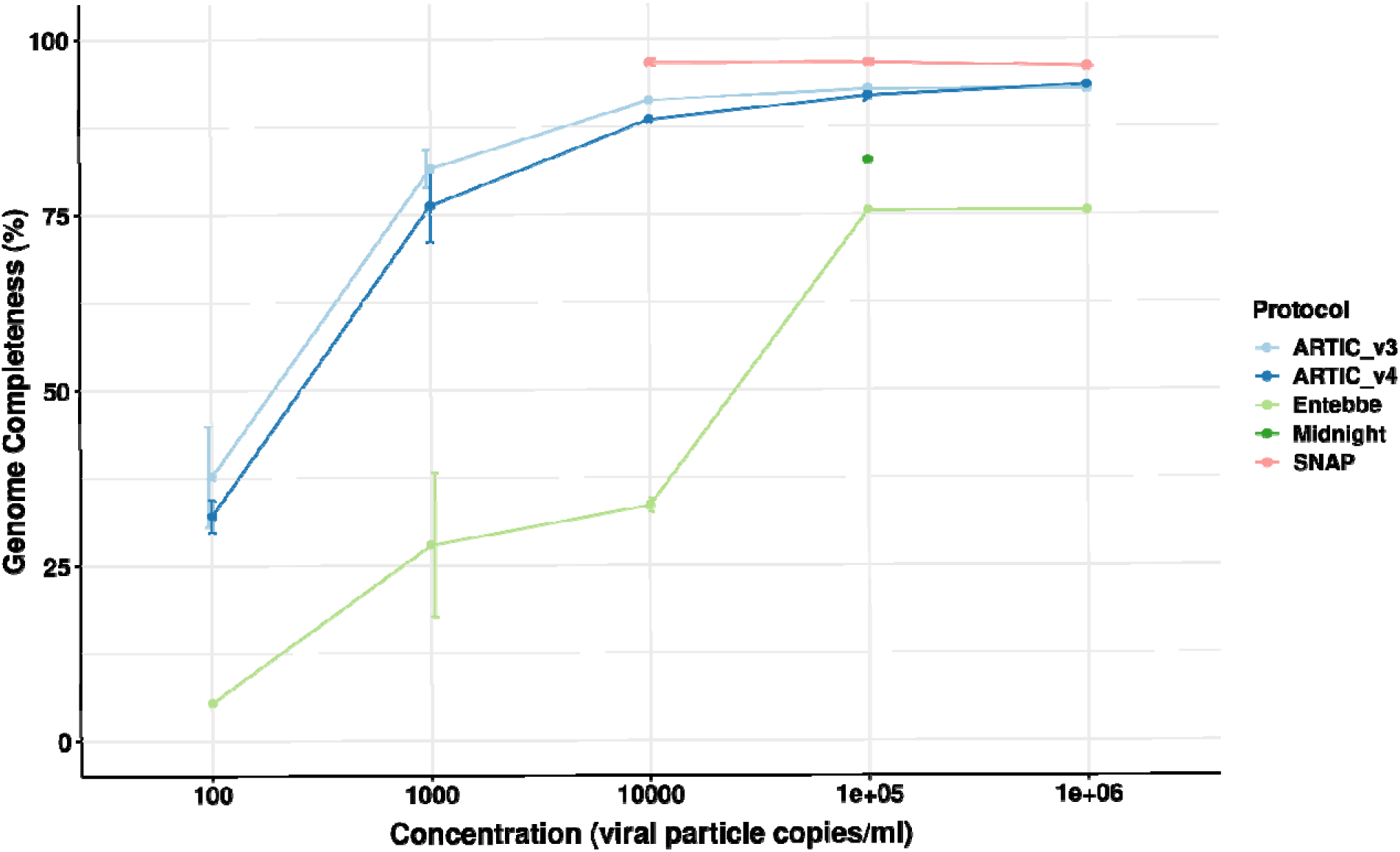
Genome completeness assessment using synthetic SARS-CoV-2 genome. The Twist Synthetic SARS-CoV-2 RNA Control 1 (102019, Twist Bioscience, CA, USA) representing wildtype SARS-CoV-2 was used as template at different log-serial diluted concentrations. cDNA used for all protocols was prepared according to the ARTIC protocol while PCR amplification followed protocol specifications. We used ARTIC v4 protocol here instead of v4.1. All data was analysed using ARTIC pipeline. Genome completeness was calculated as the percentage of the fully reconstructed genome (without gaps represented by ‘N’s). Errors bars represent standard deviation from the mean of three technical replicates.

### ARTIC protocols show highest PCR amplicon yield with cell culture SARS-CoV-2

We obtained cell culture wildtype SARS-CoV-2 (B.1) and five different variants namely, alpha, beta, gamma, delta, and omicron (B.1.1.7, B.1.351, P.1, B.1.617.2, and BA.1, respectively). The concentration of all samples was determined using digital droplet PCR and then normalized to 1000 particle/µL and finally log serial diluted to 0.1 particle/µL. Initially, we processed all samples using ARTIC v4 protocol and assessed the yield following PCR amplification (Supplementary Figure 3). With the exception of B.1.617.2 variant, samples showed similar yields with average post PCR amplicon concentrations not statistically different (One-way ANOVA p=0.386 for 100 particles/µL, p=0.687 for 10 particles/µL, p=0.06 for 1 particles/µL, and p=0.251 for 0.1 particles/µL). We then selected the 100, 1, and 0.1 viral particles/µL dilutions to represent samples with high, medium, and low viral loads, respectively. These dilutions were selected since they yielded an average of 115 ng/µL, 40 ng/µL, and 10 ng/µL (Supplementary Figure 3) which is comparable to the 1^st^ quartile (84 ng/µL), 2^nd^ quartile (31 ng/µL) and 3^rd^ quartile (8.7 ng/µL) yields observed across our clinical samples, and thus, could simulate samples at high, medium, and low viral titres, respectively.

Samples were processed with six different protocols: ARTIC v3, ARCTIC v4.1, Qiaseq, SNAP, Midnight, and Entebbe. We compared the yield of amplicons following PCR amplification across the different protocols, variants, and virus titres. The ARTIC protocols had the highest yield with version 4.1 having a slightly higher yield than version 3 except for BA.1 variant (Supplementary Figure 4). The SNAP and Qiaseq protocols had the least yields (Table 3). We sequenced all samples on the PromethION and subsampled 100, 1000, 5000, 10000, 20000, 40000, 80000, 100000, and 200000 reads from each sample and used the ARTIC pipeline to process these reads and reconstruct the genome. From each reconstructed genome, we assessed the percentage genome completeness, ability to call the correct lineage, and primer drop out.

**Table 3:** Average post-PCR amplicon yield per protocol. The mean PCR amplicon yield (ng) for the wildtype and five SARS-CoV-2 variants tested was computed for each viral titre level.

|  | **Sample viral titre (particle/μL)** |  |  |
| --- | --- | --- | --- |
|  | **100** | **1** | **0.1** |
| ARTIC_v4.1 | 109 | 37 | 14.8 |
| ARTIC_v3 | 105 | 31.2 | 9.35 |
| Entebbe | 65.2 | 9.58 | 4.2 |
| Midnight | 56.7 | 8.19 | 1.81 |
| SNAP | 6.32 | 3.21 | 1.7 |
| Qiaseq | 9.1 | 0.23 | 0.113 |

### ARTIC protocols show best overall genome completeness with cell culture SARS-CoV-2

We measured genome completeness as the percentage of the genome without ‘N’s or gaps. Overall, ARTIC protocols performed the best (Figure 3). For example, among the high viral titre samples (100 particles/µL), the ARTIC protocols had the least number of N’s followed by Entebbe protocol followed by Midnight protocol. The ARTIC v4.1 protocol required only 40,000 reads to reach maximum genome completeness of 99.58% across all variants except for B.1.351 which required 80,000 reads and only reached a maximum of 99.57% genome completeness. Interestingly, among high viral titre samples, the ARTIC v4.1 protocol performed worst with the wildtype virus where the maximum genome completeness reached was 98.75% even with 734,000 reads.

**Figure 3:**
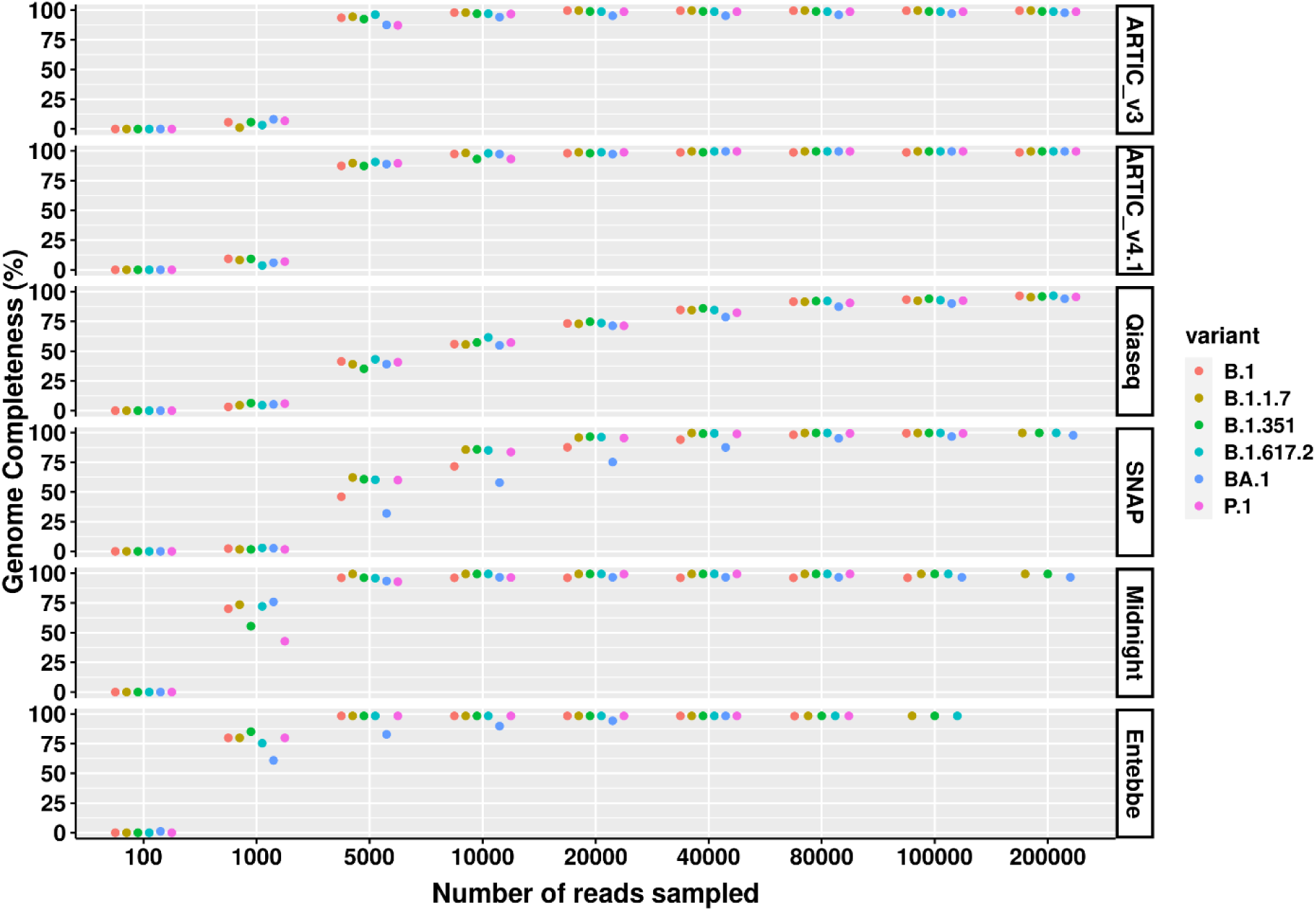
SARS-CoV-2 genome completeness comparison across high viral titre samples and protocols. Wildtype SARS-CoV-2 and five cell culture variants were processed for sequencing following six different protocols; ARTIC v3, ARTIC v4.1, Qiaseq, SNAP, Midnight, and Entebbe. Prepared cDNA libraries were sequenced on the PromethION and the data analysed using ARTIC pipeline to reconstruct the genomes using a set of randomly sub-sampled reads. At each set of sampled reads genome completeness was computed as the percentage of the fully reconstructed genome (without gaps represented by ‘N’s). Samples used here were the normalised and serial dilutions at 100 viral particles/µL.

The Entebbe protocol performed best in terms of number of reads needed to reach protocol-specific maximum genome completeness. Among high viral titre samples, only 5,000 reads were required to reach the maximum coverage obtained with this protocol, 98.33%, except for BA.1 which required 40,000 reads to reach this coverage.

The ARTIC v3 reached maximum genome completeness of 99.60% for B.1.1.7 using 20,000 reads. The variants P.1 and B.1.617.2 attained 99.60% genome completeness with 731,000 and 875,000 reads, respectively. ARTIC V3 protocol performed worst with BA.1 variant which only reached 98.66% genome completeness with 655,000 reads.

The Midnight protocol attained 99.36% genome completeness for P.1, B.1.617.2, B.1.351, and B.1.1.7 which required at least 40,000, 10,000, 10,000, and 5,000 reads, respectively. The Midnight protocol performed worst with wildtype B.1 and BA.1 which only reached 96.03 and 96.60% genome completeness with 158,000 and 233,000 reads, respectively.

The SNAP protocol reached 99.66% genome completeness with B.1.1.7, B.1.617.2, and B.1.351 variants all of which required at least 200,000 reads. With 469,000 reads, SNAP reached 99.75% genome completeness with B.1.617.2. This was the highest genome completeness of all protocols assessed. For B.1, P.1, and BA.1, the maximum genome completeness reached was 99.5, 99.28, and 97.70% using 23,000, 180,000, and 200,000 reads respectively.

The Qiaseq protocol reached a maximum of 98.04% genome completeness with the P.1 variant at 614,000 reads. Among the other samples, the maximum genome completeness reached for B.1, BA.1, B.1.1.7, B.1.351, and B.1.617.2 were 97.90, 96.11, 97.76, 96.17, and 97.14% with 0.6, 0.5, 0.8, 0.5, and 1.4 million reads, respectively.

Among medium viral load samples (1 particles/µL), we did not have enough reads to compare all protocols. Nevertheless, the ARTIC v4.1 performed best in comparison to ARTIC V3, Midnight, and Entebbe where we had comparable reads of up to 10,000 reads (Supplementary Figure 5). At 10,000 reads, ARTIC V3, Midnight, and Entebbe showed an average of 78.99, 71.58, and 71.28% genome completeness compared to 84.49% genome completeness attained with the ARTIC v4.1 protocol.

Among low viral titre samples (0.1 particles/µL), we only had enough reads from ARTIC v3 and ARTIC v4.1 protocols. ARTIC v4.1 showed 0.3 % higher genome completeness across all lineages at 100,000 reads compared to ARTIC V3 (74.51% versus 74.15%, respectively, Supplementary Figure 6).

### Long-read protocols require least reads to call correct lineage

We also assessed the number of reads needed to call the correct lineage across all lineages tested. Among high viral load samples, the Entebbe protocol required the least number of reads to correctly call all lineages tested, 1000 reads (Figure 4). This was followed by Midnight protocol which required 5000 reads to correctly call all lineages. Surprisingly, ARTIC v3 only needed 40,000 reads to correctly call all lineages while the ARTIC v4.1 needed at least 200,000 reads to correctly call BA.1. The SNAP protocol failed to correctly call BA.1 even with 200,000 reads whereas the Qiaseq protocol showed highest sensitivity to B.1.17 and B.1.351 lineages only calling all lineages correctly with 200,000 reads.

**Figure 4:**
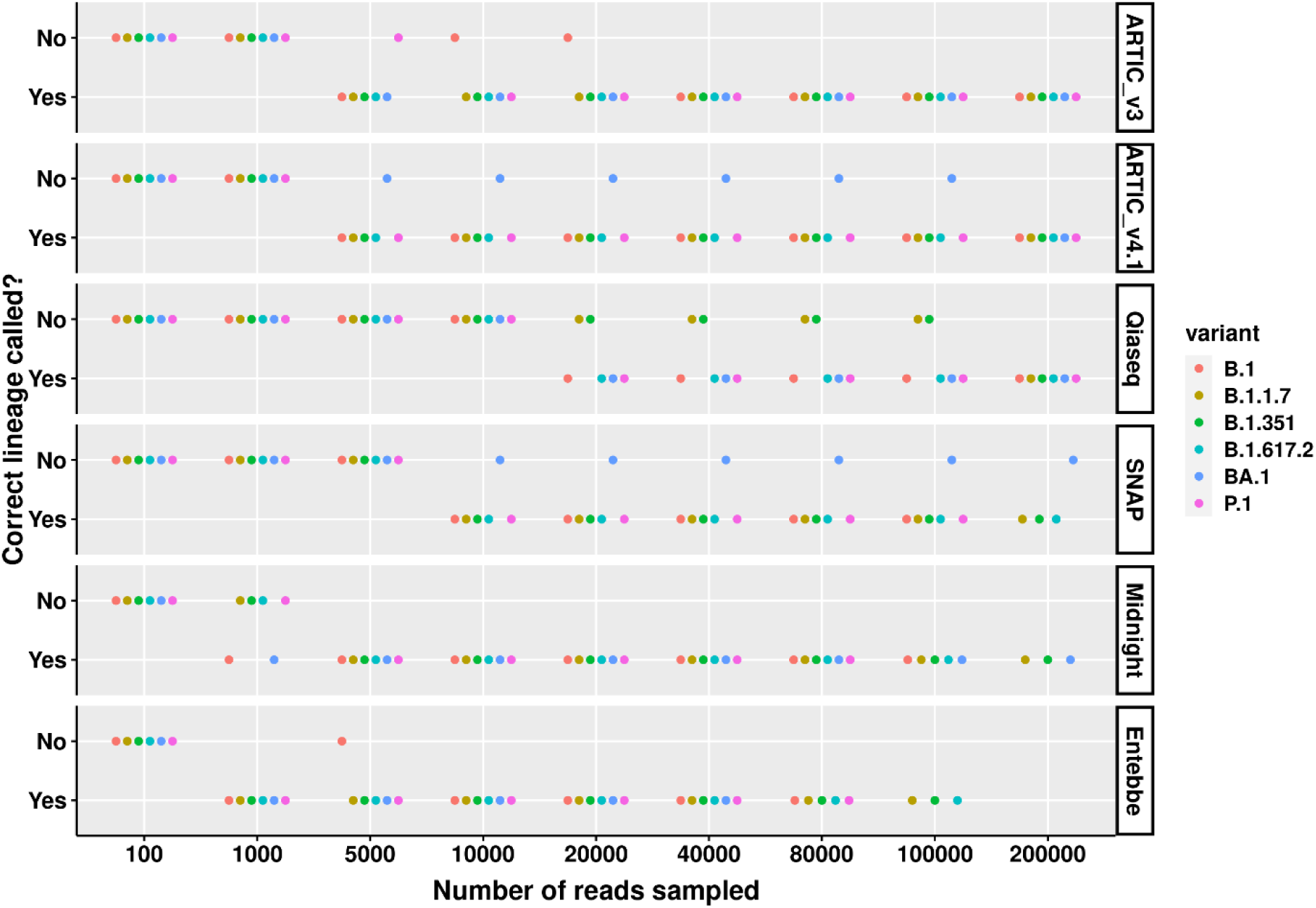
Correct SARS-CoV-2 lineage calling comparison across high viral titre variants and protocols. Wildtype SARS-CoV-2 and five cell culture variant samples were processed for sequencing following six different protocols; ARTIC v3, ARTIC v4.1, Qiaseq, SNAP, Midnight, and Entebbe. Prepared cDNA libraries were sequenced on the PromethION and the data analysed using ARTIC pipeline to reconstruct the genomes using a set of randomly sub-sampled reads. At each set of sampled reads the Pangolin pipeline (O’Toole et al., 2021) was used to assign Pango lineages (Rambaut et al., 2020) to the reconstructed genome. The figure shows whether the reconstructed genome allowed for the correct lineage to be assigned or not. Samples used here were the normalised and serial dilutions at 100 viral particles/µL.

Among medium viral load samples, the ARTIC v4.1 performed best, requiring only 5000 reads to correctly call 6 lineages tested (Supplementary Figure 7). ARTIC v3 required at least 80000 reads to call all lineages correctly. At 40000 reads, the Midnight protocol failed to correctly call B.1 and B.1.617.2 while at 20,000 reads the Entebbe protocol failed to correctly call P.1 and B.1.617.2.

Among low viral load samples, the ARTIC v3 and ARTIC v4.1 could be compared directly at 100,000 reads (Supplementary Figure 8). At this level, ARTIC v4.1 performed better only failing to return the correct lineage calls for B.1.351 and P.1 compared to ARTIC v3 which only returned correct calls for B.1 and BA.1. The Midnight protocol and Entebbe protocols, both of which are long-read protocols did not yield enough reads to evaluate them properly. Nevertheless, both protocols called BA.1 lineage correctly with only 1000 reads.

### Genome coverage and primer dropout

We assessed genome coverage for the different protocols across different lineages of SARS-CoV-2 after combining all reads to simulate a high viral load sample. Overall, the long-read protocols (Entebbe and Midnight) showed the least primer dropout with the Entebbe protocol performing better than Midnight protocol (Supplementary Figure 9 and Supplementary Figure 10). The Entebbe protocol is the only protocol that did not have any primer fail to reach a minimum threshold of 20X coverage required to call a consensus sequence by the ARTIC pipeline (primer dropout). The Midnight protocol showed sensitivity to BA.1 and the wildtype virus with two primers failing to reach 20X coverage. The ARTIC v3 protocol had reduced performance against B.1.351 and BA.1 with one and four primers, respectively failing to reach 20X coverage even when others reached 1000X coverage (Supplementary Figure 11). The ARTIC v4.1 protocol only showed one primer failing to reach 20X coverage with the wildtype virus. For all other variants, deep sequencing to achieve at least 100X average depth remedied poor primer performance (Supplementary Figure 12). SNAP and Qiaseq protocols have overlapping primers in a single tube which makes it difficult to assign reads to a single primer. Overall, these protocols achieved similar depth as the rest of the protocols although many primers showed reduced sensitivity to all viruses used (Supplementary Figure 13 and Supplementary Figure 14, respectively). This reduced sensitivity is however, compensated for by overlapping primers such that only a single region showed significant dropout for the SNAP protocol on BA.1 variant and none for Qiaseq protocol. We also assessed evenness of genome coverage measured as percentage coefficient of variation across protocol, variants, and subsampled number of reads (Supplementary Figure 15). The ARTIC v4.1 protocol showed the lowest CoV averaging ∼70% across all variants and subsampled reads, performing worst with BA.1 which averaged ∼85%. This was followed by ARTIC v3 protocol which averaged 72% across all variants and subsampled reads. The long-read protocols Midnight and Entebbe performed similarly with overall CoV average of 85% while the commercial protocols SNAP and Qiaseq performed worst with overall CoV of 86 and 103%, respectively.

### Our custom protocol drastically improves over ARTIC protocol

Given the poor performance of all protocols on samples with medium and low viral load titres, we sought a method that would increase sensitivity of SARS-CoV-2 detection. We tweaked a recently published method called Rolling Circle Amplification to Concatemeric Consensus (R2C2) (Volden et al., 2018) and termed it ARTIC-Amp (for amplified ARTIC) as it leverages the ARTIC v4.1 protocol and complements it with rolling circle amplification. In a “proof of principle” experiment, we selected four primer-sets from the ARTIC v4.1 primers that target four genes namely, N, ORF7a, ORF1a, and Spike, respectively and added extra tag sequences to allow for circularisation via Gibson assembly. To this effect, we also amplified a 330 bp ‘splint’ with Lambda virus DNA as the template using primers carrying the same tag sequences (Supplementary Figure 16A). Next, we optimised primer annealing and extension temperatures and found that unlike ARTIC v4.1 primers that had the highest yield at 64.3 °C, the ARTIC-Amp modified primers had highest yield at 65.6 °C (Supplementary Figure 16B). Using this optimised annealing temperature, we completed the ARTIC-Amp protocol and generated products with peak length of 8.9 kb (Supplementary Figure 16C). We compared the ARTIC v4.1 PCR amplicon yields and ARTIC-Amp final yields following rolling circle amplification (RCA) (Supplementary Figure 17). We noted that whereas the ARTIC v4.1 yields dropped with reducing viral titres, ARTIC-Amp yields remained relatively stable across viral titres except for BA.1 (BA.1_100) sample (Supplementary Figure 17). On average, the ARTIC-Amp protocol resulted in a 40X amplification of yields among the low viral titre samples. For example, for both B.1.617.2 and BA.1 low viral titre samples that were done in triplicate, the average amplicon yields increased from 1.0 and 1.3 ng/µL to 40 and 44 ng/µL, for ARTIC v4.1 and ARTIC-Amp protocols, respectively (Supplementary Figure 18). Sequencing of these products on the PromethION resulted in products with median length of 805 bp (Supplementary Figure 19). The number of reads generated with the ARTIC-Amp was evenly distributed across samples compared to the reads generated from ARTIC v4.1 which corresponded to viral titre (Supplementary Figure 20).

We then assessed the number of times each original amplicon was amplified by rolling circle amplification of circularised product. The number of repeats identified in each sequence had a median of 1 and a mean of 2. The distribution of the number of repeats in each read is shown in Supplementary Figure 21. We looked at 12394 sequences for which we could get a consensus following error correction. The alignment identity of these sequences increased by 6%, from 91% without consensus correction to 97 % following R2C2-mediated consensus correction (Supplementary Figure 23 and Supplementary Figure 22). Finally, we compared genome coverage between ARTIC v4.1 and ARTIC-Amp in the B.1.617.2 low viral titre samples. We noted that whereas two targeted regions were missed in two out the three low viral titre replicates for the ARTIC v4.1 protocol no region was missed in any of the three low viral titre replicates for the ARTIC-Amp protocol (Figure 5).

**Figure 5:**
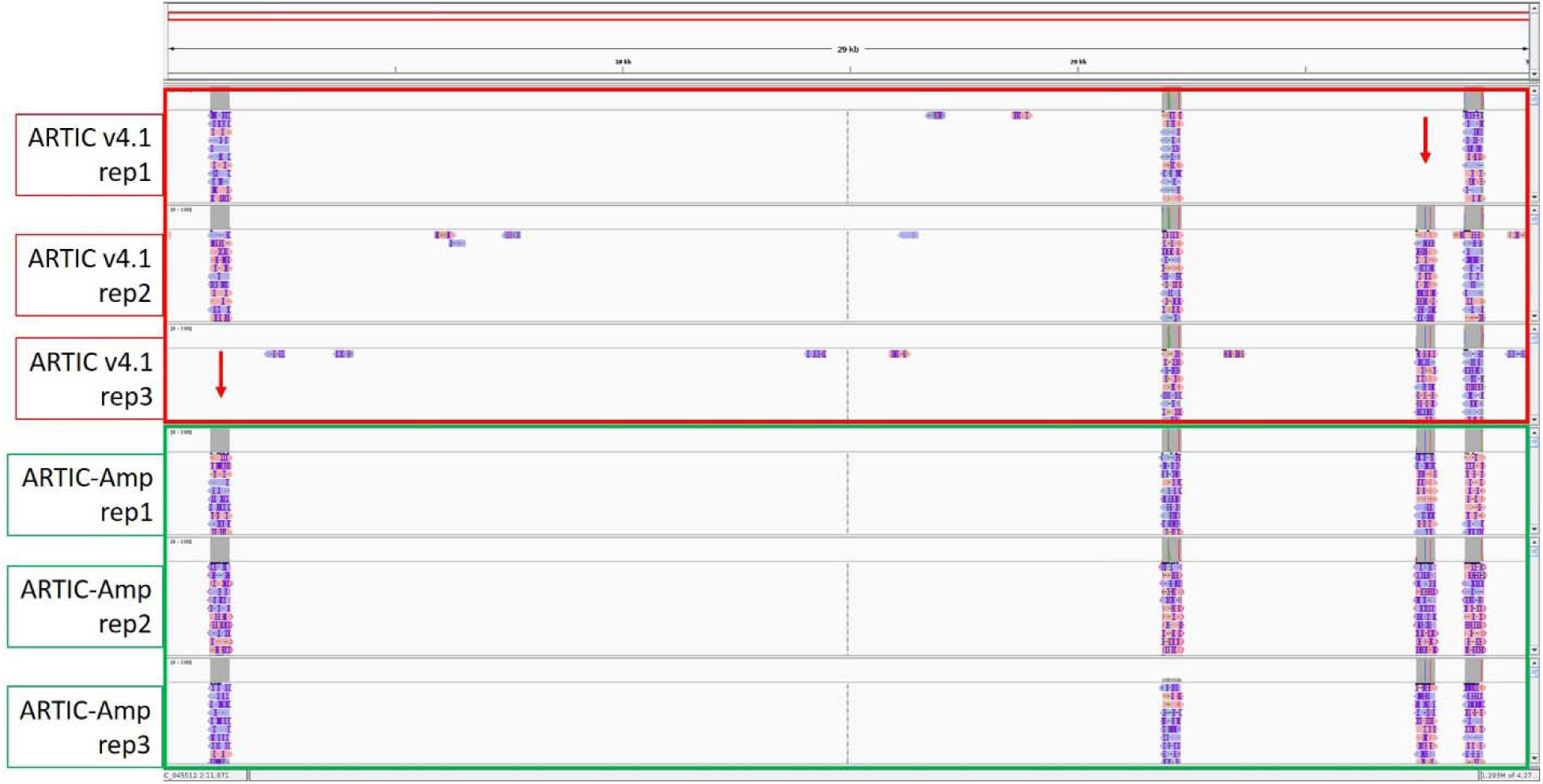
Comparison of genome coverage between ARTIC v4.1 and ARTIC-Amp protocols. Four primer sets targeting regions in four SARS-CoV-2 genes: N, ORF7a, ORF1a, and Spike, respectively were used to prepare sequencing libraries either following the ARTIC v4.1 protocol or our in-house protocol termed ARTIC-Amp. The ARTIC-Amp protocol takes the final products of the ARTIC v4.1 protocol and circularises them via Gibson assembly followed by isothermal rolling circle amplification as described previously (Volden et al., 2018). Reads generated by the ARTIC-Amp protocol were processed using R2C2 pipeline (Volden et al., 2018). All reads were aligned to the genome and loaded on IGV (Robinson et al., 2011) for viewing. A screenshot is shown here. Red arrows indicate regions missed by ARTIC v4.1 protocol. No regions were missed by ARTIC-Amp protocol in this proof-of-principle experiment. Samples used here were the B.1.617.2 low viral titre (0.1 viral particles/µL) done in triplicate.

## Discussion

The COVID19 pandemic started in late 2019 and continues to rage throughout the world causing losses both in life and labour shortages due to disease burden. Several methods have emerged for high throughput whole genome sequencing of SARS-CoV-2 in clinical and wastewater samples. These methods can be categorised based on the target enrichment method used. The two most commonly employed target enrichment methods are target hybrid capture and PCR (also referred to as amplicon-based methods). Target capture uses target-specific probes while PCR uses target specific primers. The former include such methods as KAPA SARS-CoV-2 Target Enrichment Panel (Roche) and Illumina RNA Prep with Enrichment Tagmentation (Illumina) which use hybridization probes to capture SARS-CoV-2 sequences (hybrid capture). In one study, capture-based methods were found to have higher accuracy of identifying ‘within-sample variation’ compared to amplicon-based methods while amplicon-based methods had higher accuracy of identifying ‘between-sample variations’ (Xiao et al., 2020). Perhaps the most widely applied method is one developed by the ARTIC network (https://artic.network/ncov-2019) which belongs to the amplicon-based methods. The primers used in this method have undergone at least three changes to cope with SARS-CoV-2 evolution. In the current work we evaluated version 3 and 4.1 of the ARTIC primers. Further, we evaluated the Swift Normalase Amplicon Panel for SARS-CoV-2 (SNAP, Swift Bioscience), QIAseq DIRECT SARS-CoV-2 (Qiaseq, Qiagen), Midnight, and Entebbe protocols on post-PCR amplicon yield, genome completeness, correct lineage calling, and primer dropout. The ARTIC protocols were included because of their ease of use and wide adoptability, SNAP and Qiaseq are commercial protocols while Midnight and Entebbe are academic research protocols. ARTIC, SNAP, and Qiaseq are short read protocols with reads averaging 430, 350, and 350 bp, respectively while Midnight and Entebbe are long-read approaches with reads averaging 1200 and 1800 bp, respectively. The Entebbe protocol was also included because its reverse transcription step uses target-specific primers unlike all the other protocols that use random hexamers. However, we realized that in our hands, using random hexamers doubled the PCR amplicon yield (Supplementary Figure 24). We, therefore, performed cDNA synthesis using random hexamers for the Entebbe protocol.

Initially, we used a synthetic genome diluted at different viral copy concentrations. The synthetic genome was composed of 5 kb non-overlapping fragments. The non-overlapping fragments, however, create a unique situation since SARS-CoV-2 positive samples would be expected to have either full genome and/or randomly sheared and overlapping fragments. We evaluated post-PCR concentration of amplicons because in our experience this generally correlates positively with genome completeness. We observed the highest amount of PCR amplicons using the ARTIC v4 protocol compared to all other protocols suggesting a higher PCR efficiency with this protocol. We utilized the ARTIC v4 primers for this experiment since the version 4.1, updated to address mutations in BA.1, was not yet available and would not be envisaged to improve yields of this wildtype synthetic virus. The SNAP protocol yielded the lowest concentrations of all protocols. Although this could probably be due to the lower number of PCR cycles recommended for this protocol compared to other protocols (28 versus 36 cycles, respectively), increasing PCR cycles to 36 in a later experiment did not increase yields. Nevertheless, the SNAP protocol showed the highest genome completeness at 1×10^4^, 1×10^5^, and 1×10^6^ copies/mL where we had enough reads to compare across protocols. This is probably due to two factors. 1) The SNAP protocol has 344 primers which is by far the highest compared to 98 for ARTIC v3, 99 for ARTIC v4, 20 for Entebbe, and 29 for Midnight. 2) The synthetic genome having been composed of non-overlapping segments. The high number of primers in the SNAP protocol probably had better ability to cover the ends of the fragments than the other protocols could thus resulting in better genome completeness. The SNAP protocol, therefore, should perform well with highly degraded samples. However, the high number of primers also means higher sensitivity of the primers to mutations (Borcard et al., 2022; Kuchinski et al., 2022; Rosenthal et al., 2022). In fact, the SNAP primers underwent at least one round of changes in response to viral evolution. The ARTIC protocols were the second in achieving highest genome completeness with insignificant differences between version 3 and 4. The two long read protocols, Entebbe and Midnight performed worst probably due to the lower number of primers. We only had enough reads to evaluate the Midnight protocol at 1×10^5^ copies/mL where it performed better than Entebbe protocol.

Next, we used cell culture wildtype and variant SARS-CoV-2. The samples were quantified using digital droplet PCR (ddPCR) and then normalized in an attempt to have a comparable number of viral particles in each sample. Samples were then serial-diluted and then processed with ARTIC v4 protocol to determine the amplicon concentration. We have found that post-PCR amplicon concentration is indicative of genome completeness such that samples with 50 ng/µL and above have a positive correlation with genome completeness and yield the highest coverage. We then used three dilutions of cell culture SARS-CoV-2; 100 particles/µL to reflect samples with high viral titre, 1 particle/µL to reflect samples with medium viral titres, and 0.1 particle/µL to reflect samples with low viral titres. When processed with ARTIC v4.1 in triplicate, our normalisation was successful with no significant differences between yields for the wildtype and the four variants tested at 100 viral particles/µL and 0.1 particles/µL. The B.1.617.2 variant did not seem to be as well normalised as the other samples which caused the samples at 1 particle/µL to be significantly different. The normalization however, worked well given that the post-PCR amplicon yield was comparable between the four ddPCR quantified samples and the Twist biosciences synthetic virus suggesting that they contained similar viral particles (Supplementary Figure 25).

The main goal of whole genome SARS-CoV-2 sequencing is to obtain the whole sequence of the genome which provides the entirety of mutations acquired by the virus. Therefore, genome completeness, as determined by the percentage of the genome that is assembled without gaps represented as ‘N’ is critical (Chiara et al., 2021). Genome completeness is positively correlated with sample viral titre such that samples with low viral titre have reduced genome completeness. We thus, evaluated genome completeness in samples with high (100 particles/µL), medium (1 particles/µL), and low viral titres (0.1 particles/µL). Among samples with high viral titres, the SNAP protocol achieved the highest genome completeness of 99.75 % with 469,000 reads except for B.1.617.2 variant. The SNAP protocol performed worst with BA.1 variant. Noteworthy, we evaluated an earlier version of the SNAP protocol which did not include revisions to address virus evolution. The SNAP protocol thus, performed best with genome completeness but required very high sequencing depth. Choi et al (Choi et al., 2022) reported genome completeness of 99.78 % with SNAP although no lineage information was reported. They further used ddPCR and reported that SNAP required a minimum of 10.5 copies/µL to achieve >95 % genome completeness.

Among all protocols tested, it was the ARTIC v4.1 protocol that achieved highest coverage with the lowest number of reads. Further, the ARTIC v4.1 protocol performed relatively well across variants. The ARTIC protocol showed highest coverage across high and low viral titre samples in other studies (Charre et al., 2020; Nasir et al., 2020). The Qiaseq protocol achieved modest genome completeness attaining a maximum of only 98.04 % which was the lowest among the protocols we tested. Noteworthy, we prepared the cDNA as described for ARTIC protocol and only followed the Qiaseq protocol for PCR amplification. Further, the Qiaseq and SNAP protocols were developed primarily with Illumina sequencing in mind although we did not see any technical barrier to sequencing libraries generated by these protocols with the PromethION.

Given high viral load titres, the long-read protocols showed least primer dropout. This is likely due to the lower number of primers employed in these protocols (20 and 29 primers, respectively). We find that the Entebbe primers are more suitable than Midnight primers in this case. However, we only tested the primers yielding 1200 bp amplicons from the Midnight protocol and did not test their other counterparts which yield 1500 bp and 2000 bp amplicons (N. E. Freed et al., 2020). It is possible that these longer amplicon Midnight primers perform comparably to the Entebbe primers which yield 1800 bp amplicons. Although we did not specifically assess genome coverage at medium and low viral loads, all evidence points towards the ARTIC protocol performing better than other protocols. In this work, the ARTIC protocols generated enough libraries even from medium and low viral load samples. Further, Liu et al, reported highest genome coverage from ARTIC-based protocol among seven different protocols they tested (Liu et al., 2021). We further found that the ARTIC v4.1 protocol had the best evenness of genome coverage measured as coefficient of variation at 70%. This was also reported by Liu et al. who found that ARTIC-based protocol performed best out of the seven they tested on uniformity of genome coverage (Liu et al., 2021). Liu et al, used 1 million Illumina reads and found the ARTIC-based protocol to have a coefficient of variation of 100%. Using Nanopore reads, we find that ARTIC protocols have an average 70% which might be related to the length of reads. Therefore, overall, we find that the ARTIC v4.1 protocol outperforms other protocols in terms of low primer dropout and evenness of genome coverage.

Because of the relatively poor results obtained with medium and low viral titre samples across all protocols we sought to design a method that would improve sensitivity of amplification of SARS-CoV-2 and thus improve genome completeness and correct lineage calling among medium and low viral titre samples. We adapted a previously employed method called R2C2 (Volden et al., 2018). The authors had used this method to improve accuracy of the relatively high error-prone Nanopore reads. However, we envisaged that the isothermal multiple displacement amplification (MDA)-mediated rolling circle amplification (RCA) used in the R2C2 method would also work to increase abundance of amplicons. We called this method ARTIC-Amp (for amplified ARTIC) since it leverages the simplicity and robustness of the ARTIC protocol. Thus, samples were processed initially according to the ARTIC v4.1 protocol and then taken through another round of RCA overnight. In our proof-of-principle experiment that included only four primers targeting four different SARS-CoV-2 genes, we noticed that indeed, there is an improvement of 40X in the amount of amplicons following overnight RCA. Further, this reaction seems to proceed to its endpoint and thus results in similar amounts of material in each sample and thus normalises the amplicons eliminating the need to perform further normalisation for multiplexed samples. This normalisation further results in comparable number of reads once the samples are barcoded, multiplexed and sequenced on the same flow cell. We however, noticed that although we see a peak of 8.9 kb in the RCA products, the sequenced reads had a peak at ∼1.5 kb. Consequently, the number of repeats in each sequence was very low with a median of one. It is likely that more stringent size selection will be beneficial to increase the read lengths. We also need to devise strategies of improving circularisation efficiency. This would help to increase consensus accuracy of reads. We achieved a 6 % increase in read alignment rate, but this was for a small portion of reads that had enough repeats to create a consensus. Our main goal was to use the R2C2 method to increase sensitivity of detection. In our proof-of-principle experiment, this was achieved. As shown in Figure 5, we were able to recover all four target regions in the BA.1 low viral titre triplicate samples processed with ARTIC-Amp protocol yet two of these regions were missed in three of the same replicates processed with just ARTIC v4.1 protocol. Logistical and time reasons have hindered full exploration of this protocol but remains part of our ongoing work in the lab. We however, strongly believe that this approach can dramatically improve sensitivity of detection and be very useful for samples that routinely have low viral titres such as wastewater samples. Further, this protocol only adds USD 21 per sample over the cost of generating ARTIC v4.1 amplicons which we think is reasonable (see supplementary materials for reagent costs). Further, since amplicon-based methods have been found to outperform capture-based methods particularly in challenging samples (Xiao et al., 2020), we believe that our approach which leverages the robust amplicon-based ARTIC protocol and further complements it with isothermal amplification should provide superior sensitivity.

## Data Availability

The datasets presented in this study have been deposited to NCBI SRA with project number PRJNA887365. The individual SRR numbers for each dataset can be found in Supplementary Table 1

## Acknowledgments

We would like to thank Qiagen and Swift Biosciences for free test kits they provided some of which were used in generating data for the current work. We thank members of the McGill Genome Centre CovSeq project for their help with sequencing of samples.

## Author contribution statement

AB designed the study and conceived the ARTIC-Amp protocol, conducted the experiments, performed data analysis, and drafted the manuscript. SJR advised on study design. LF participated in the design of the study. ID and AG performed experiments and were involved in the data collection. JR oversaw the project execution. All authors read, corrected, and approved the final version of this article.

## Ethics statements

**Studies involving animal subjects:** No animals were involved in this study.

**Studies involving human subjects:** No human studies are presented in this manuscript.

**Inclusion of identifiable human data:** No potentially identifiable human images or data is presented in this study.

## Supplementary figures

**Supplementary Figure 1:**
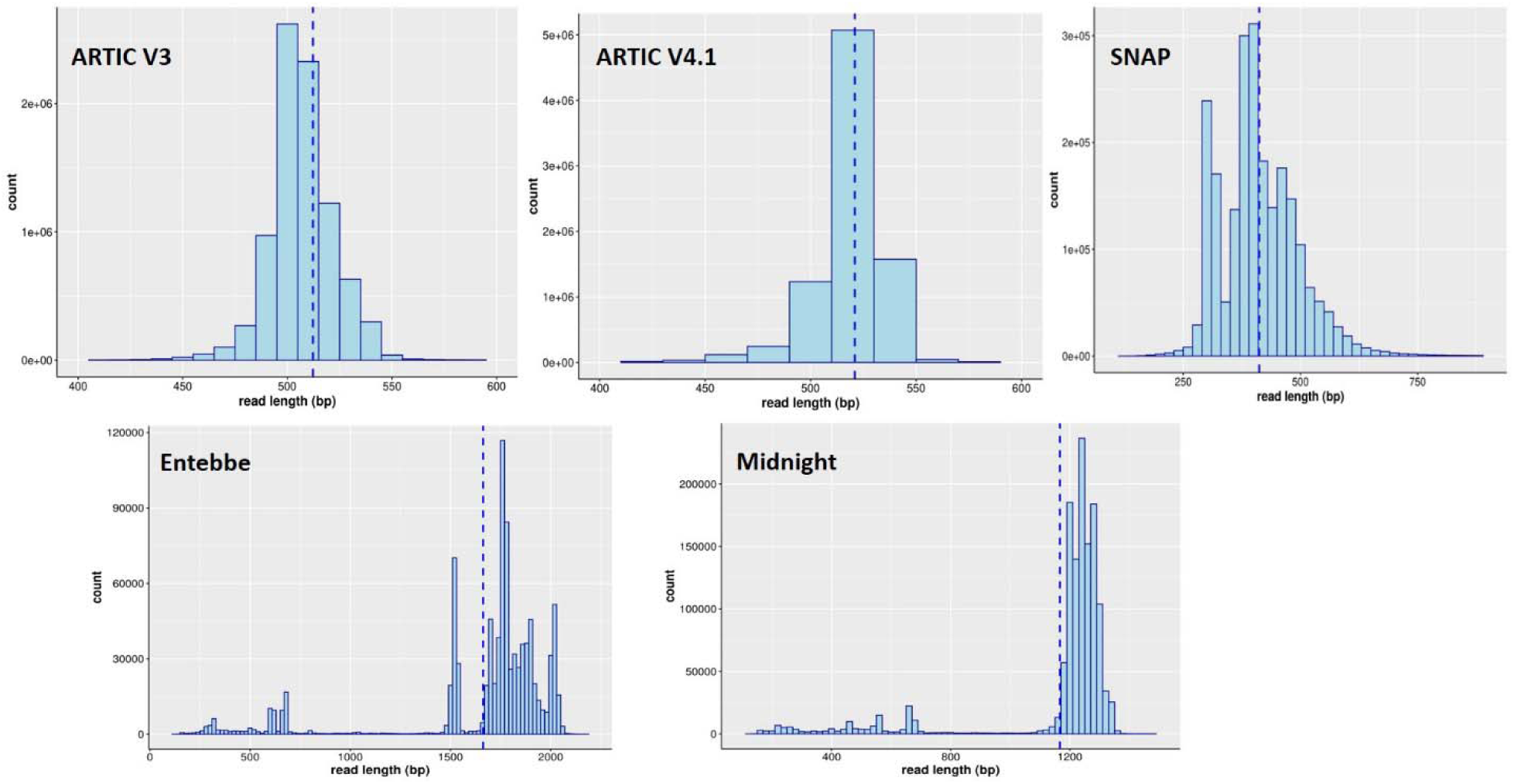
Raw read length profiles. SARS-CoV-2 samples were processed following five different protocols: ARTIC v3, ARTIC v4.1, SNAP, Entebbe, and Midgnight to generated sequencing libraries. Libraries were sequenced on the Oxford Nanopore Technologies PromethION. The profile of raw reads generated is shown here. Blue dotted line shows the median read length.

**Supplementary Figure 2:**
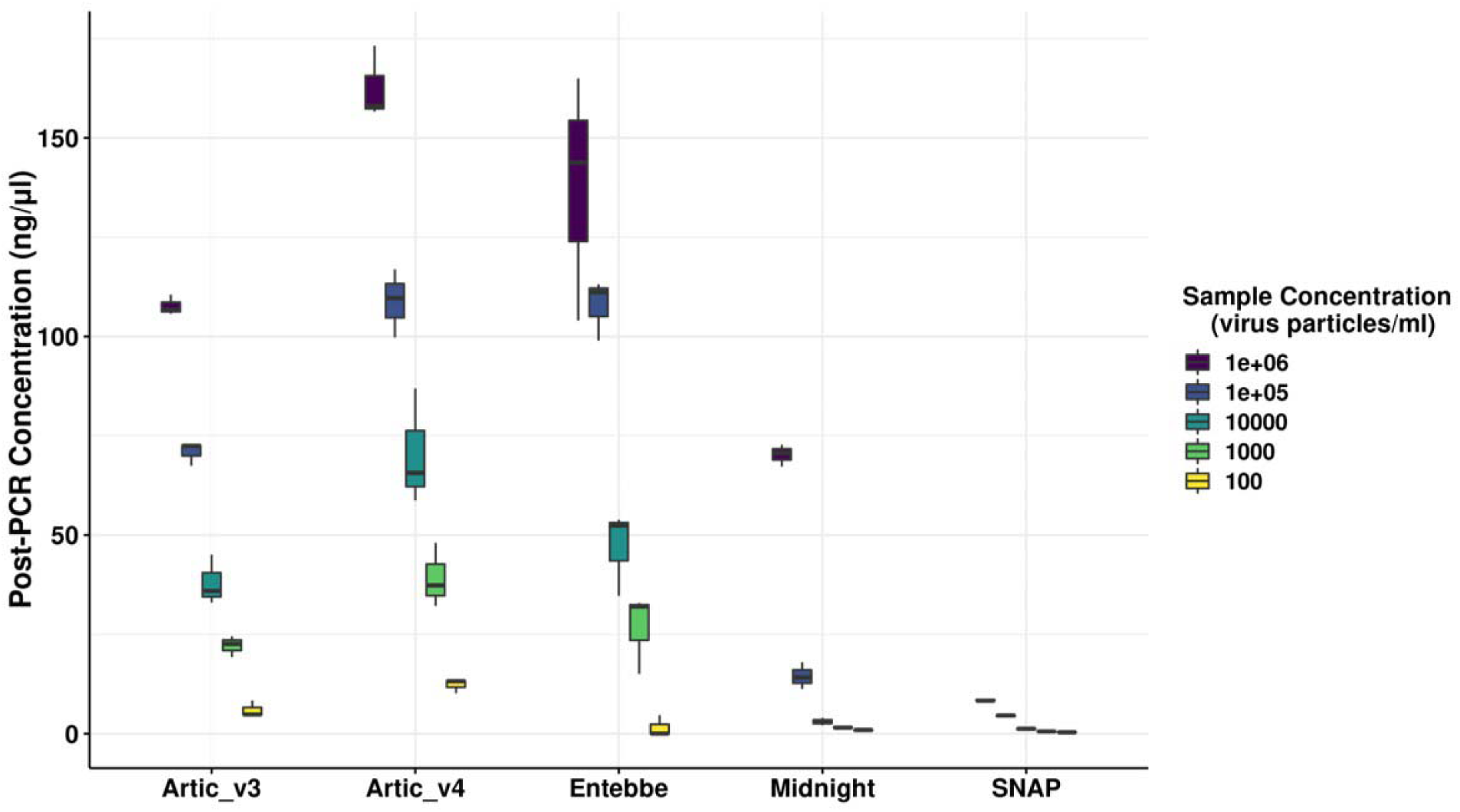
Post-PCR concentration of synthetic genome amplicons. The Twist Biosciences synthetic genome representing wildtype SARS-CoV-2 was serial diluted and the samples processed in triplicate following five different protocols: ARTIC v3, ARTIC v4, Entebbe, Midnight, and SNAP. The concentration of purified PCR amplicons was measured and shown here.

**Supplementary Figure 3:**
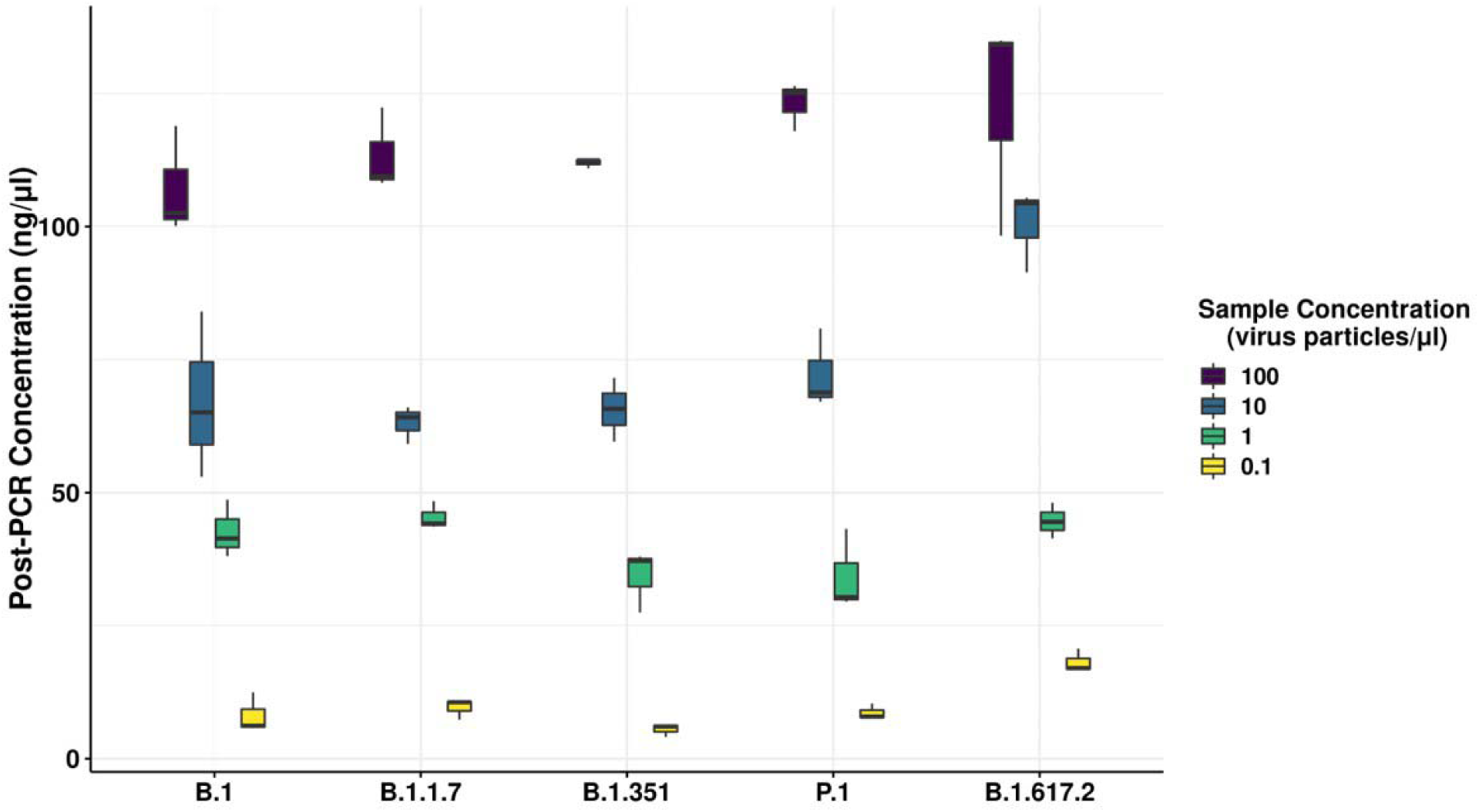
Concentration of PCR amplicons of SARS-CoV-2 cell culture variants. The concentration of wildtype and 4 different SARS-CoV-2 variant samples was determined by digital PCR followed by normalisation of the samples. The normalised samples were serial diluted and sequencing libraries prepared following ARTIC v4.1 protocol. The concentration of purified PCR products is shown.

**Supplementary Figure 4:**
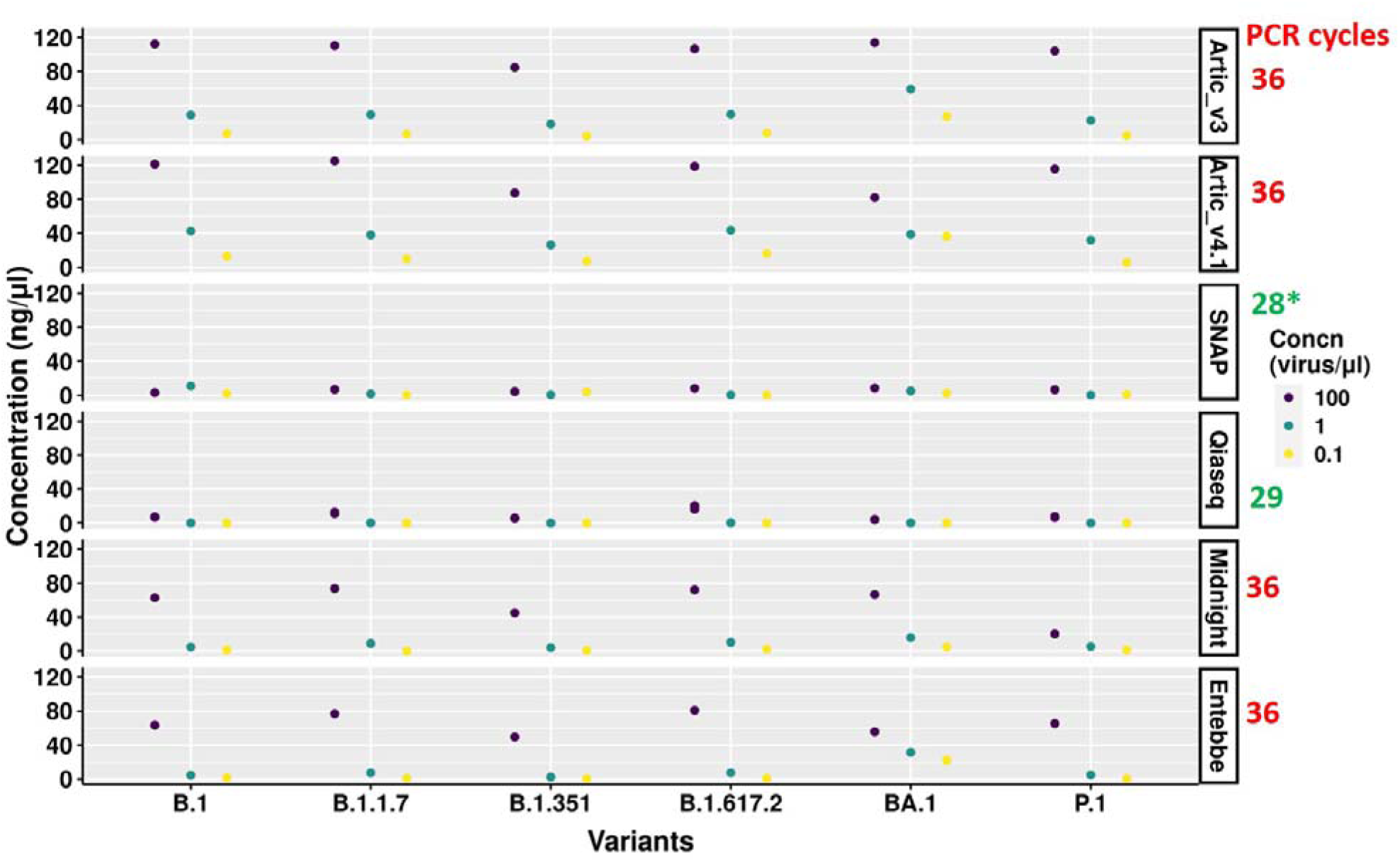
Concentration of PCR amplicons of SARS-CoV-2 cell culture variants across protocols. Normalised and serial diluted wildtype and five different SARS-CoV-2 variant samples at 3 viral titres: 100, 1, and 0.1 particles/µL were processed following six different protocols: ARTIC v3, ARTIC v4.1, Qiaseq, SNAP, Entebbe, and Midnight. The concentration of purified PCR products is shown here. The number of PCR cycles performed for each protocol is shown in red on the right of the figure. For the SNAP protocol, we performed 36 cycles for BA.1 and 28 cycles for all other samples. For Qiaseq protocol, all samples were run in triplicate and three dots representing the three samples are shown albeit that they overlap.

**Supplementary Figure 5:**
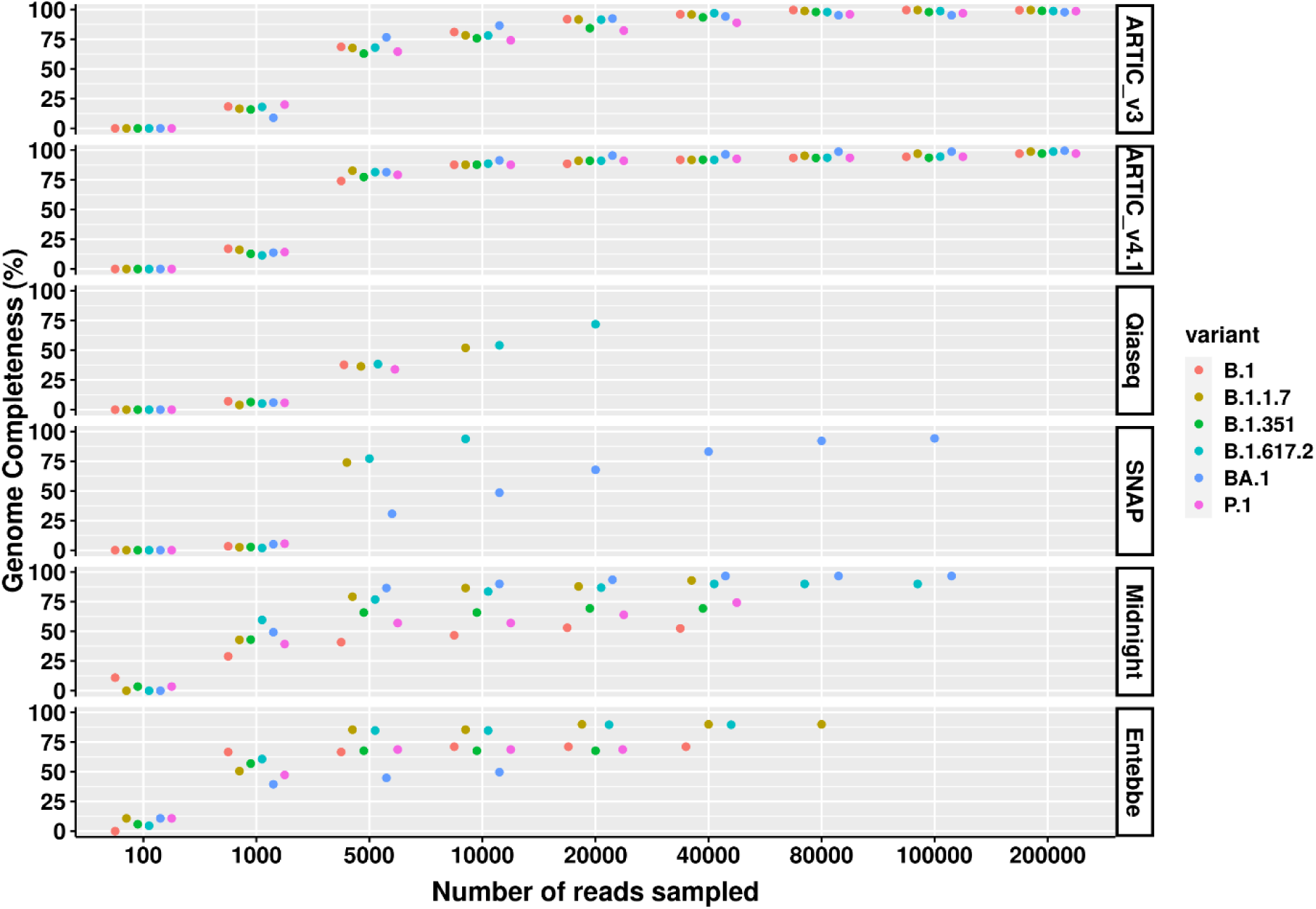
SARS-CoV-2 genome completeness comparison across medium viral titre variants and protocols. Wildtype SARS-CoV-2 and five cell culture variant samples were processed for sequencing following six different protocols; ARTIC v3, ARTIC v4.1, Qiaseq, SNAP, Midnight, and Entebbe. Prepared cDNA libraries were sequenced on the PromethION and the data analysed using ARTIC pipeline to reconstruct the genomes using a set of randomly sub-sampled reads. At each set of sampled reads the coverage of the reconstructed genome was computed as the percentage of the fully reconstructed genome (without gaps represented by ‘N’s). Samples used were the normalised and serial dilutions at 1 viral particles/µL.

**Supplementary Figure 6:**
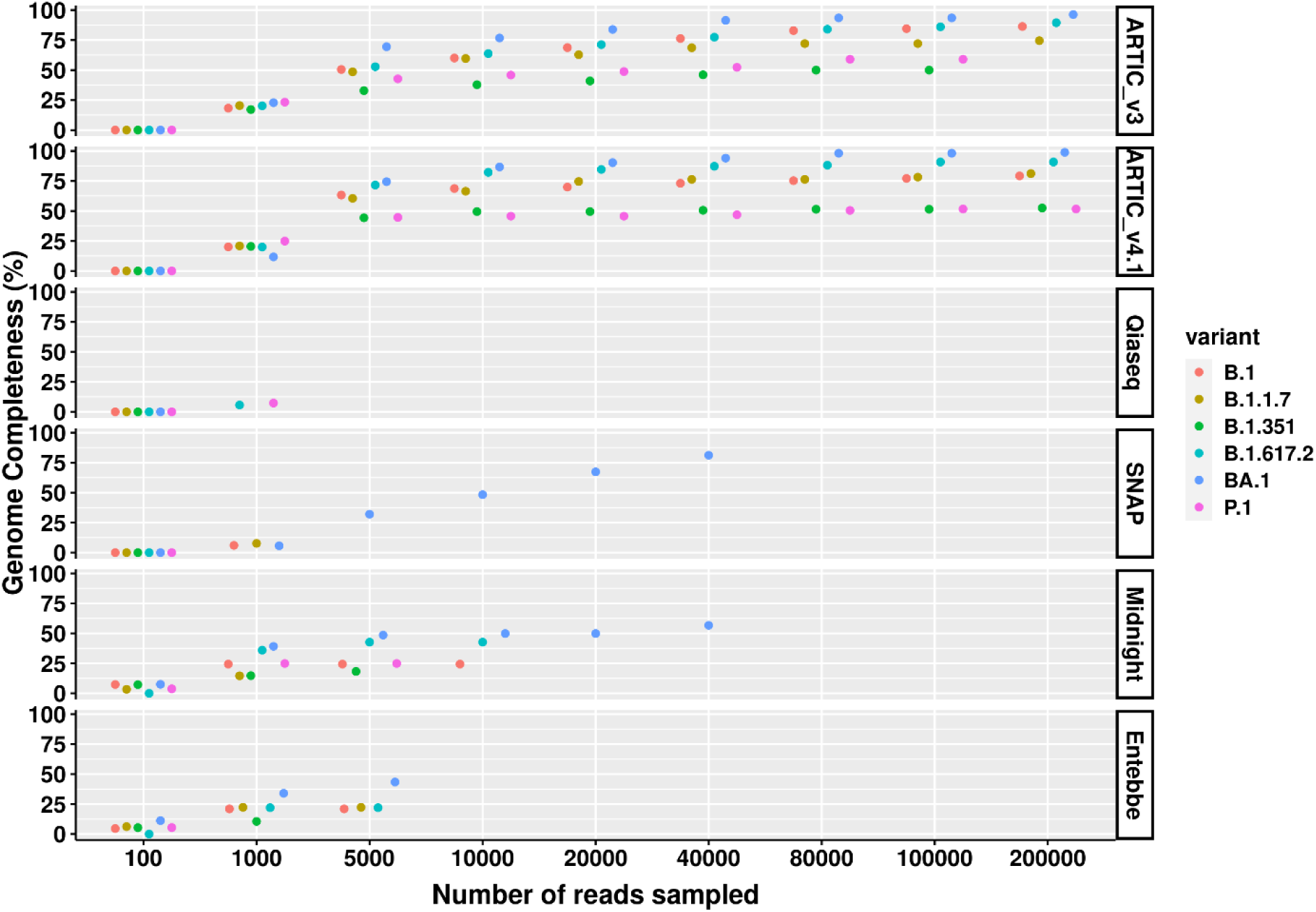
SARS-CoV-2 genome completeness comparison across low viral titre variants and protocols. Wildtype SARS-CoV-2 and five cell culture variant samples were processed for sequencing following six different protocols; ARTIC v3, ARTIC v4.1, Qiaseq, SNAP, Midnight, and Entebbe. Prepared cDNA libraries were sequenced on the PromethION and the data analysed using ARTIC pipeline to reconstruct the genomes using a set of randomly sub-sampled reads. At each set of sampled reads the coverage of the reconstructed genome was computed as the percentage of the fully reconstructed genome (without gaps represented by ‘N’s). Samples used were the normalised and serial dilutions at 0.1 viral particles/µL.

**Supplementary Figure 7:**
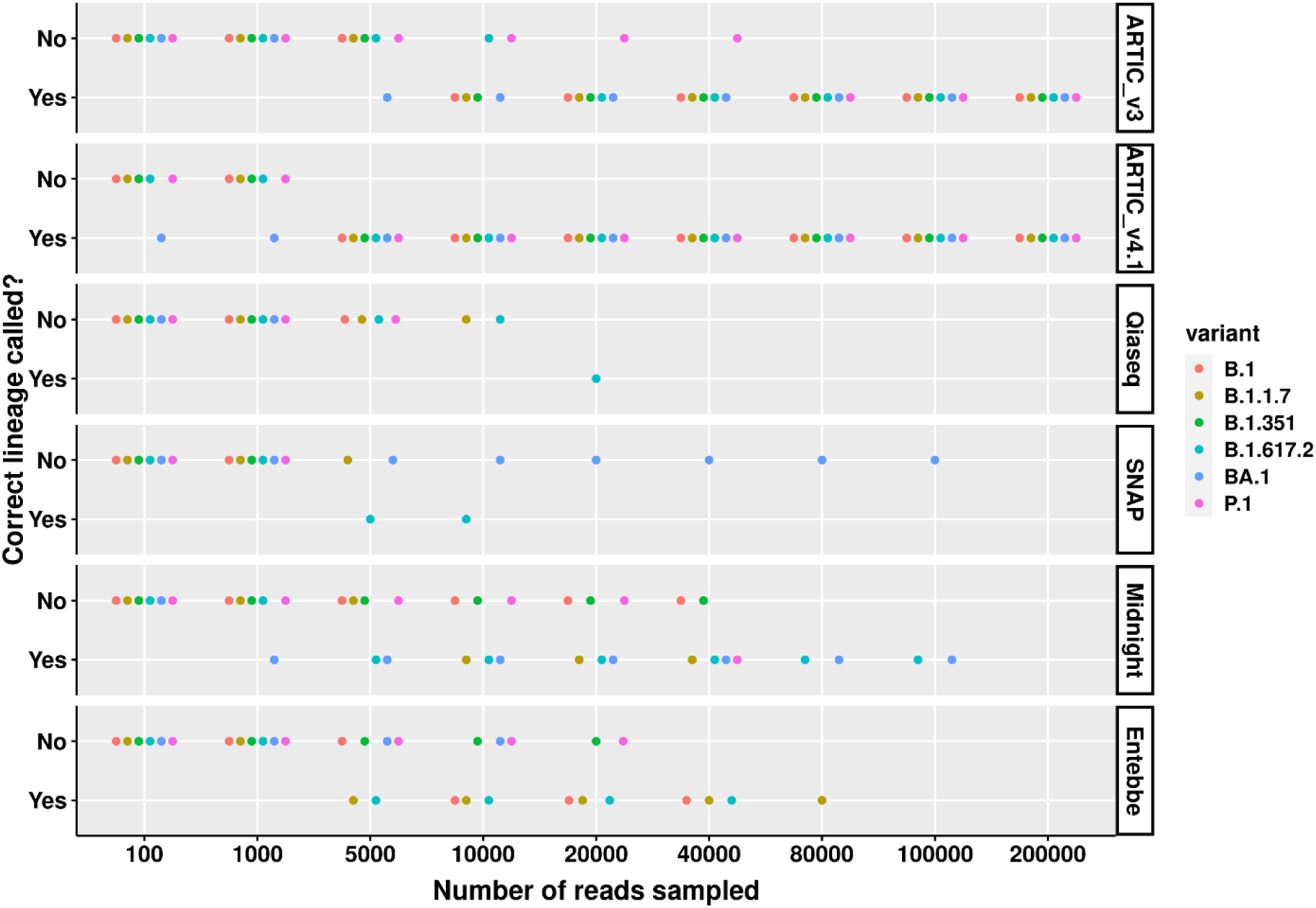
Correct SARS-CoV-2 lineage calling comparison across medium viral titre variants and protocols. Wildtype SARS-CoV-2 and five cell culture variant samples were processed for sequencing following six different protocols; ARTIC v3, ARTIC v4, Qiaseq, SNAP, Midnight, and Entebbe. Prepared cDNA libraries were sequenced on the PromethION and the data analysed using ARTIC pipeline to reconstruct the genomes using a set of randomly sub-sampled reads. At each set of sampled reads the Pangolin pipeline (O’Toole et al., 2021) was used to assign Pango lineages (Rambaut et al., 2020) to the reconstructed genome. The figure shows whether the reconstructed genome allowed for the correct lineage to be assigned or not. Samples used were the normalised and serial dilutions at 1 viral particles/µL.

**Supplementary Figure 8:**
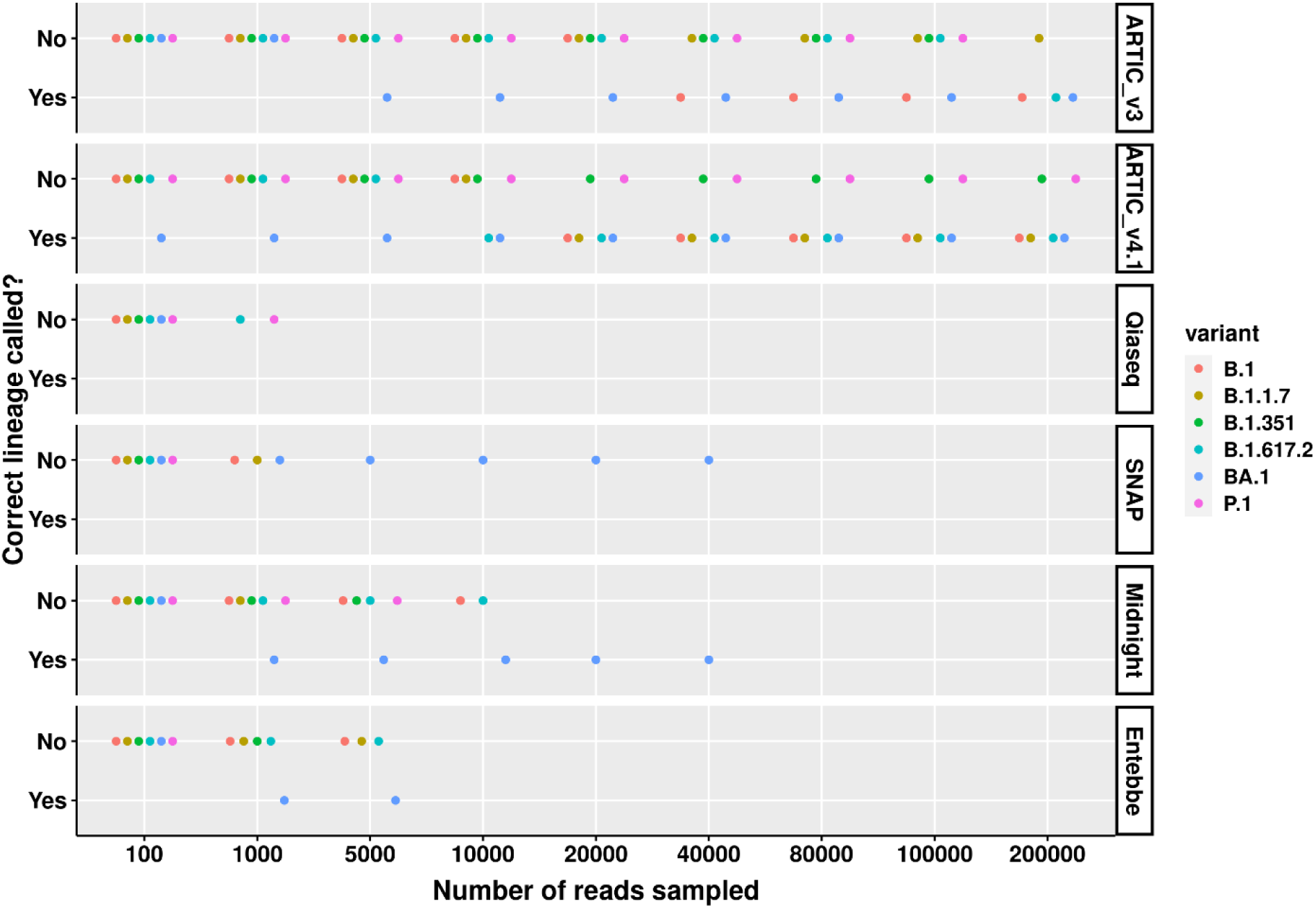
Correct SARS-CoV-2 lineage calling comparison across low viral titre variants and protocols. Wildtype SARS-CoV-2 and five cell culture variant samples were processed for sequencing following six different protocols: ARTIC v3, ARTIC v4, Qiaseq, SNAP, Midnight, and Entebbe. Prepared cDNA libraries were sequenced on the PromethION and the data analysed using ARTIC pipeline to reconstruct the genomes using a set of randomly sub-sampled reads. At each set of sampled reads the Pangolin pipeline (O’Toole et al., 2021) was used to assign PANGO lineages (Rambaut et al., 2020) to the reconstructed genome. The figure shows whether the reconstructed genome allowed for the correct lineage to be assigned or not. Samples used were the normalised and serial dilutions at 0.1 viral particles/µL.

**Supplementary Figure 9:**
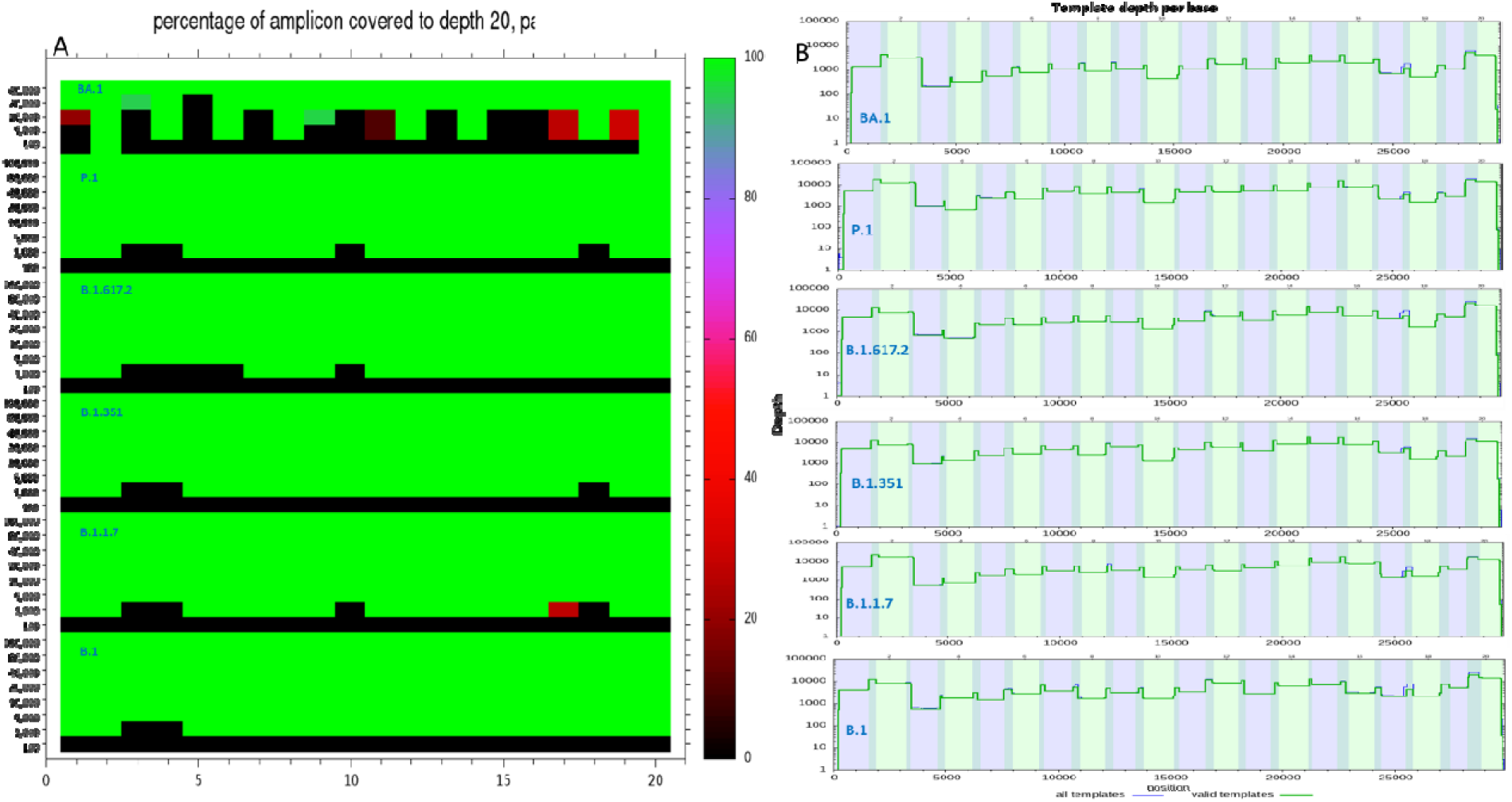
Genome coverage for Entebbe protocol. A) Heatmap showing percentage of Entebbe protocol primers covered at ≥ 20X using the number of reads subsampled and across 6 SARS-CoV-2 lineages; B.1, B.1.1.7, B.1.351, B.1.617.2, P.1, and BA.1 at high viral titres. The primer numbers are shown on the X-axis while the subsampled reads are shown on the Y-axis. B) Genome coverage depth across SARS-CoV-2 lineages using all reads (no subsampling). The depth of coverage is shown on the y-axis. The bottom x-axis shows SARS-CoV-2 genomic position while top x-axis shows the primer number. The images were generated using the “plot-ampliconstats” function of Samtools (Li et al., 2009) and bam files containing aligned reads.

**Supplementary Figure 10:**
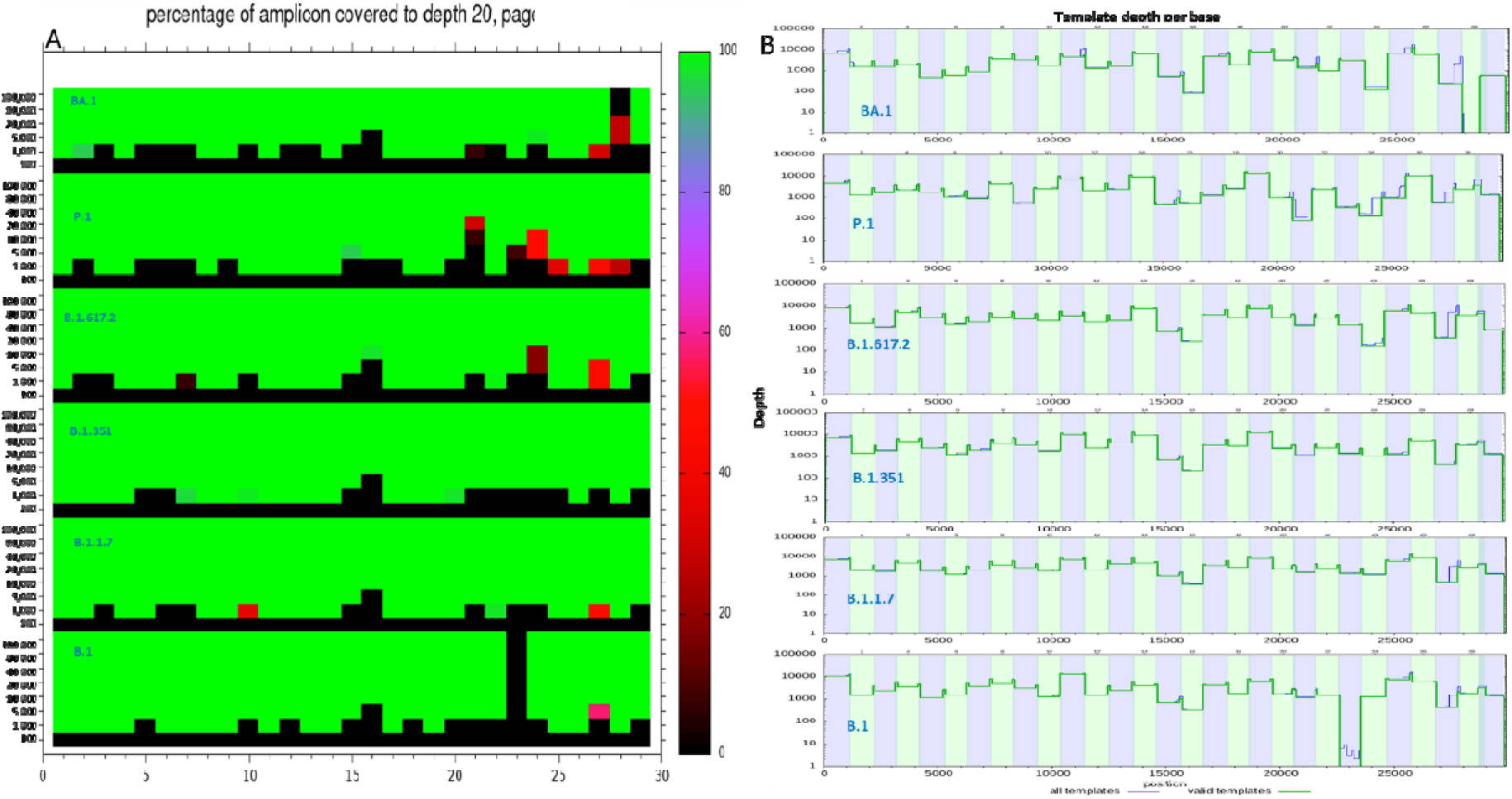
Genome coverage for Midnight protocol. Figures A and B are same as Supplementary Figure 9 except the results are from Midnight protocol.

**Supplementary Figure 11:**
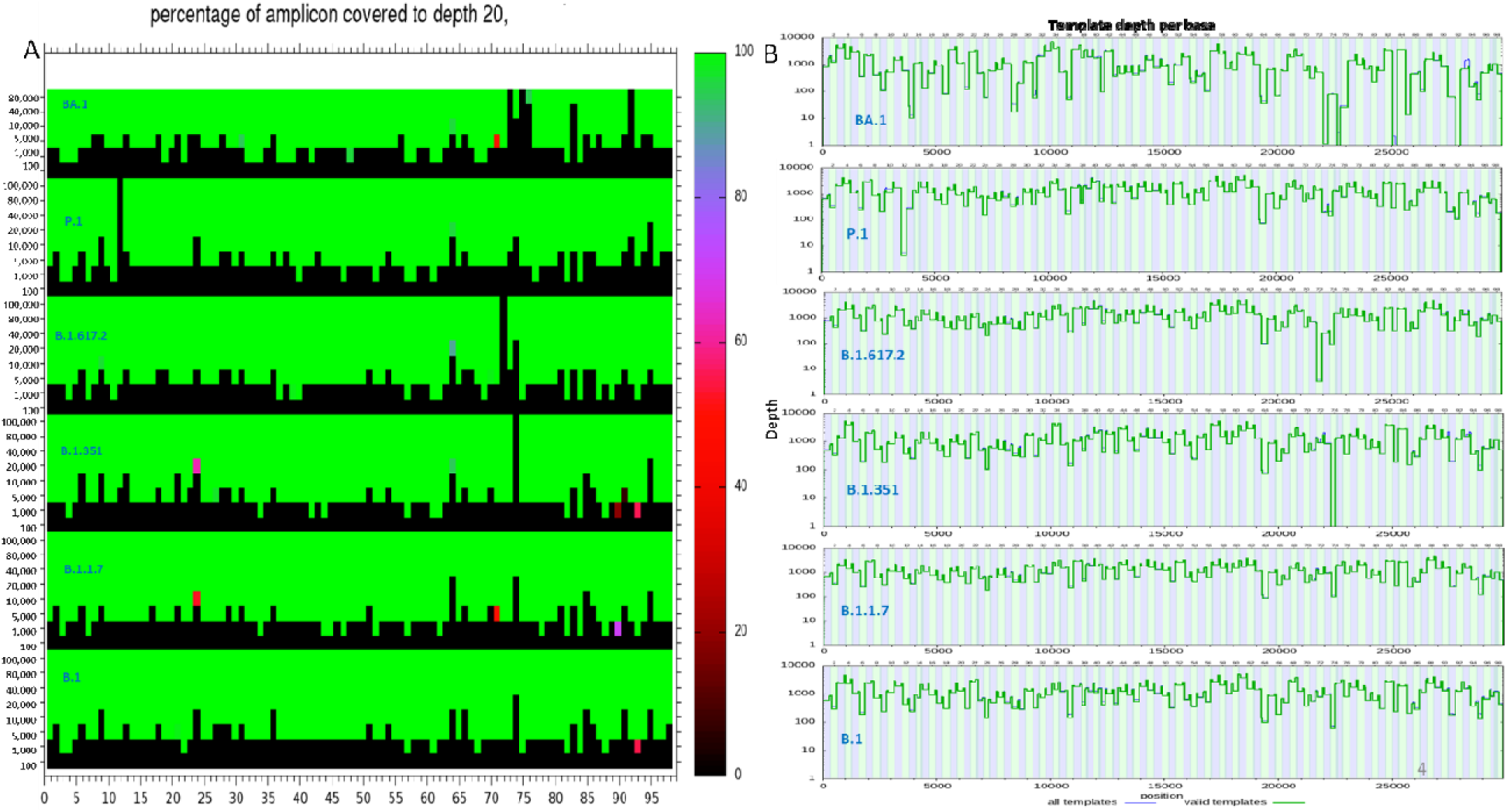
Genome coverage for ARTIC v3 protocol. Figures A and B are same as Supplementary Figure 9 except the results are from Midnight protocol.

**Supplementary Figure 12:**
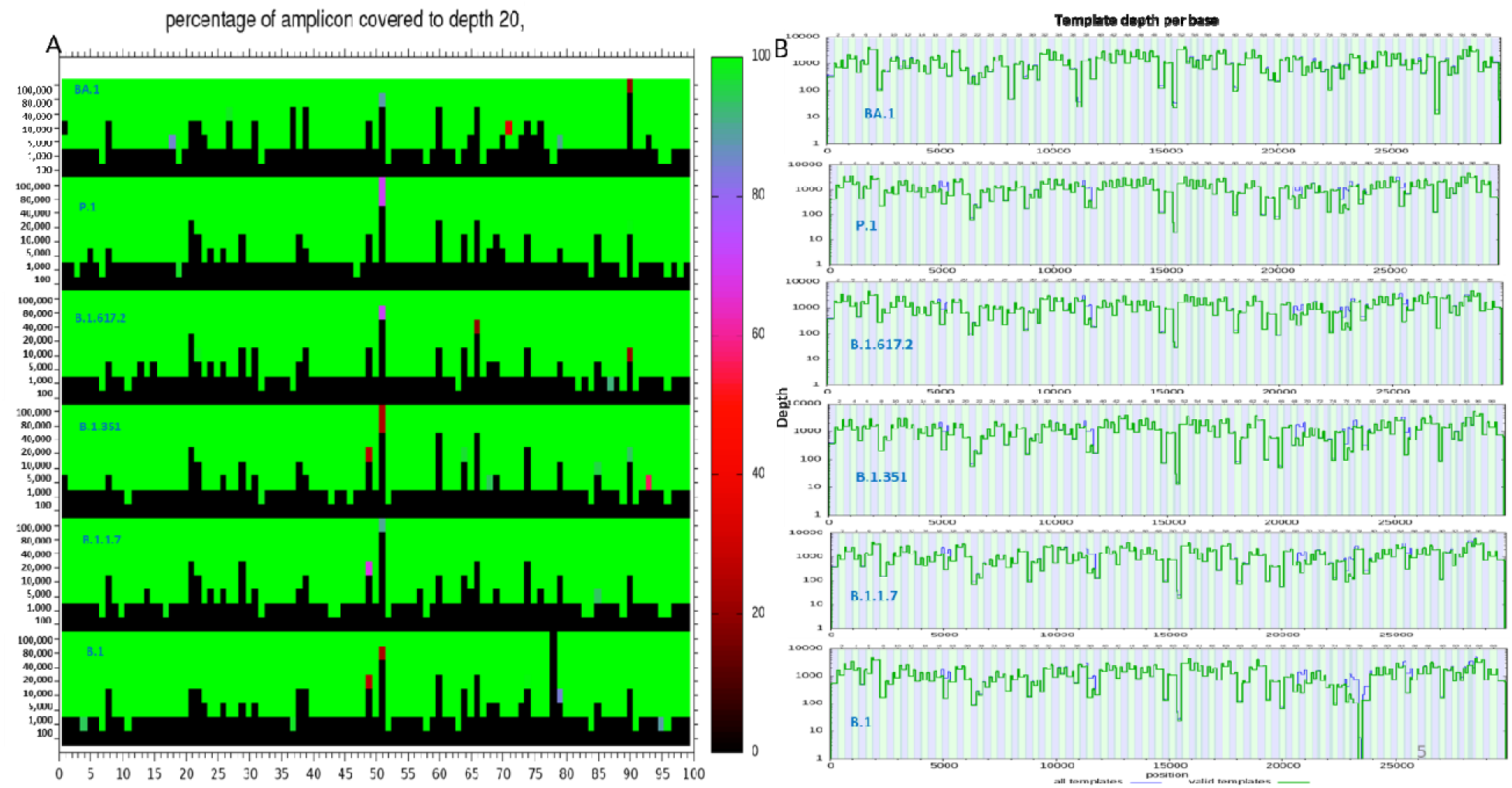
Genome coverage for ARTIC v4.1 protocol. Figures A and B are same as Supplementary Figure 9 except the results are from Midnight protocol.

**Supplementary Figure 13:**
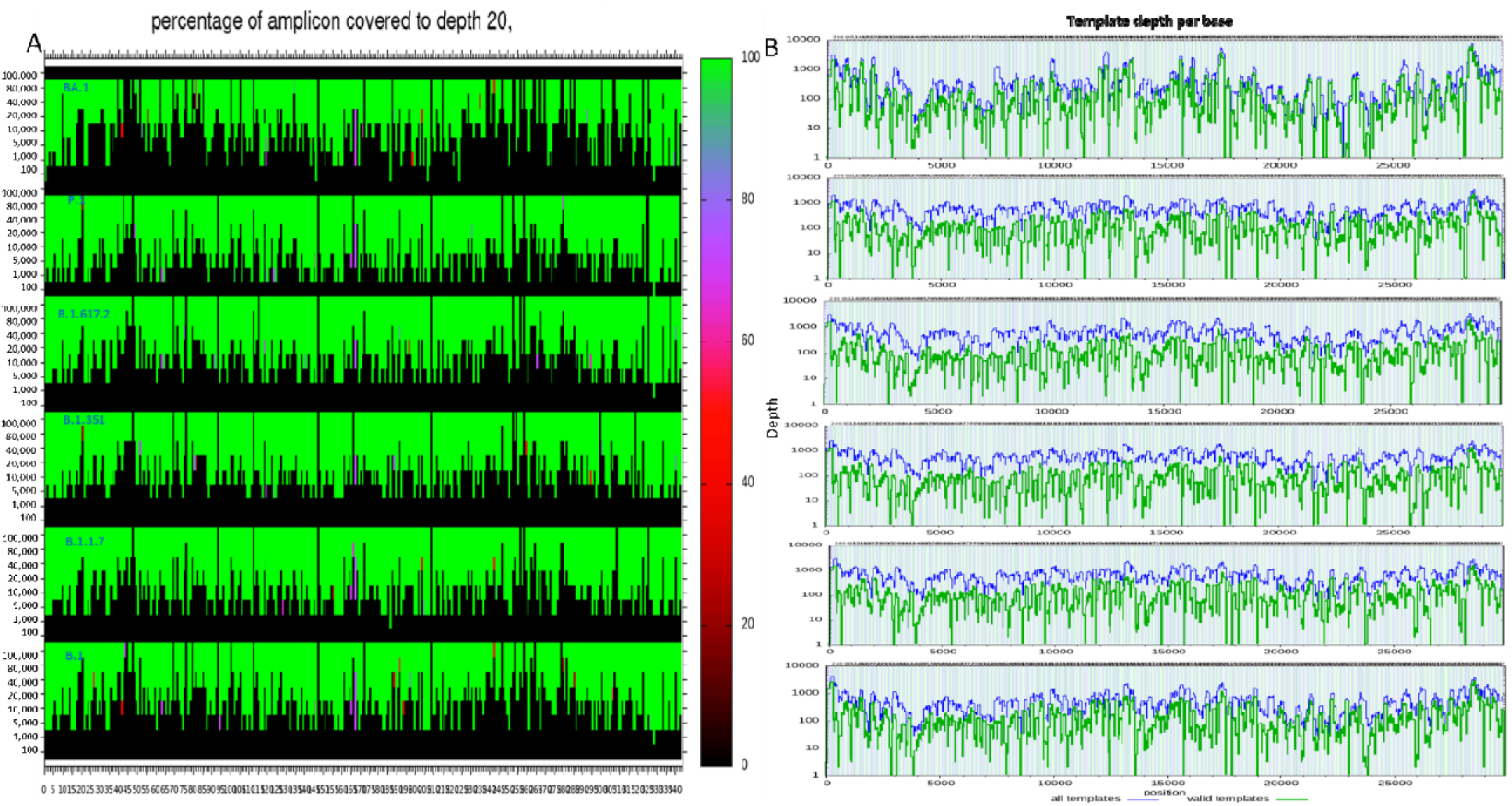
Genome coverage for SNAP protocol. Figures A and B are same as Supplementary Figure 9 except the results are from Midnight protocol.

**Supplementary Figure 14:**
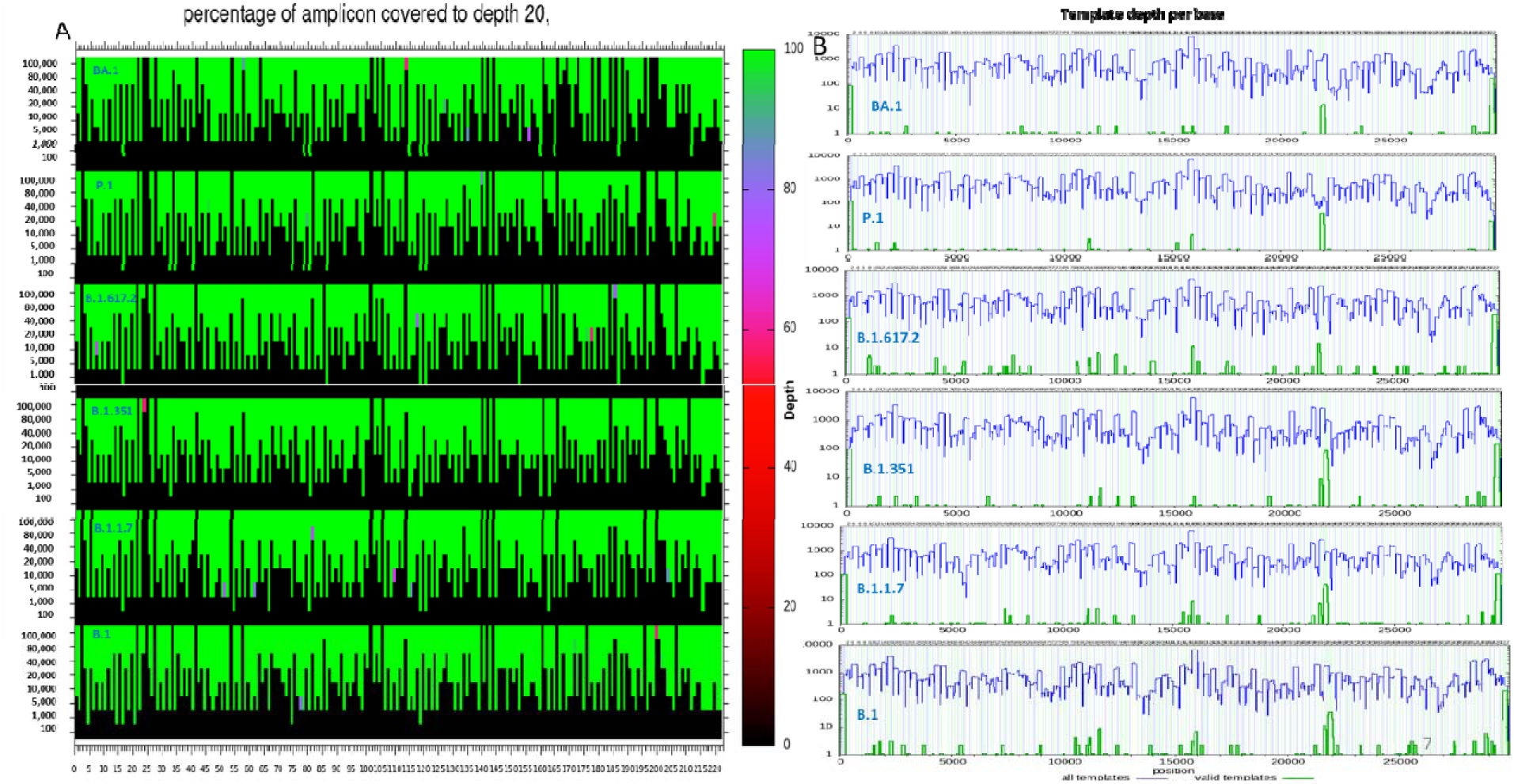
Genome coverage for Qiaseq protocol. Figures A and B are same as Supplementary Figure 9 except the results are from Midnight protocol.

**Supplementary Figure 15:**
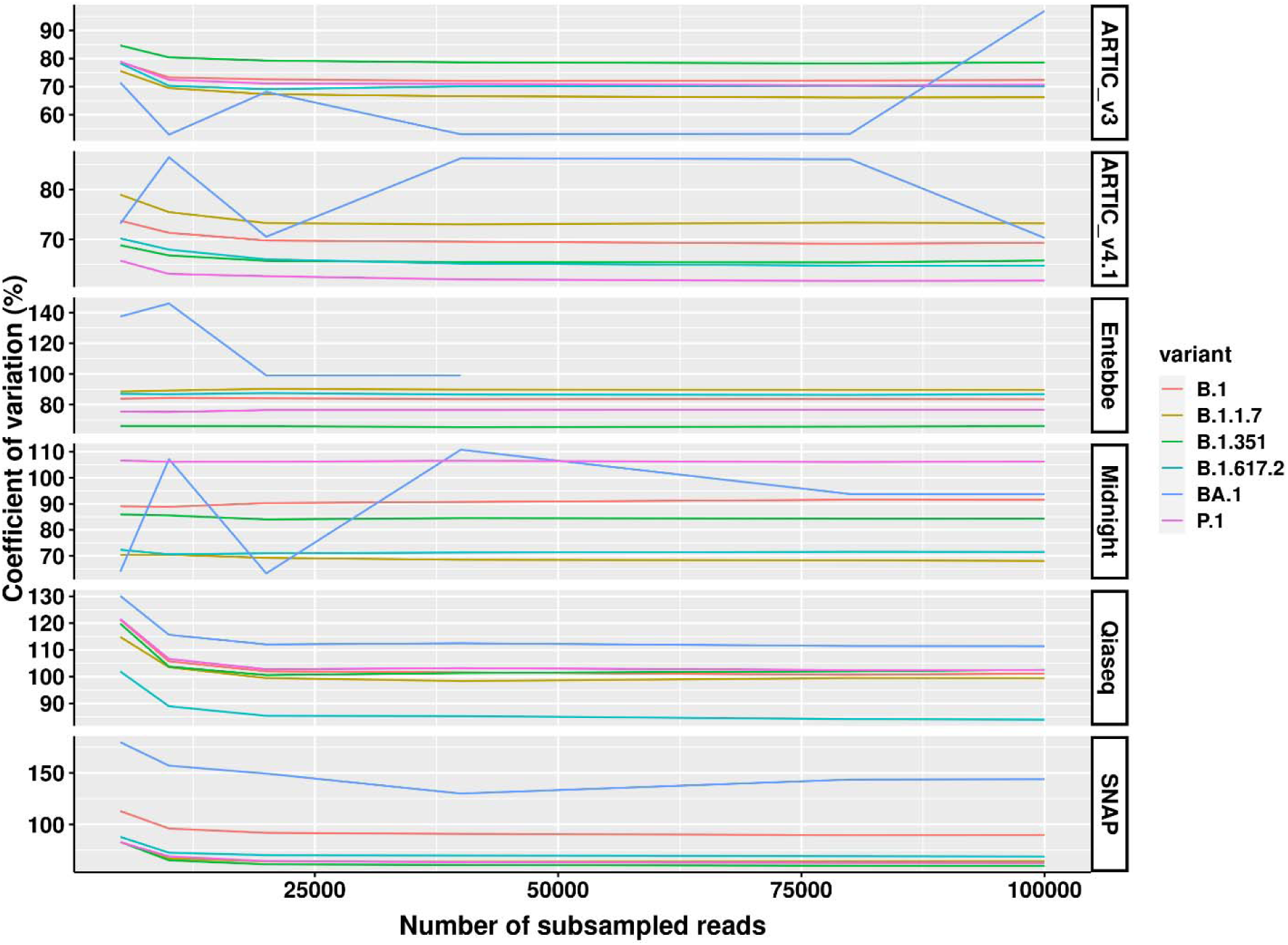
Evenness of genome coverage. Six samples containing high viral titre SARS-CoV-2 lineages namely B.1, B.1.1.7, B.1.351, B.1.617.2, P.1, and BA.1 were processed with six protocols namely ARTIC v3, ARTIC v4.1, Entebbe, Midnight, Qiaseq, and SNAP. The reads generated were subsampled and aligned to the genome and the coverage determined using the “samtools depth” function of Samtools (Li et al., 2009). The coefficient of variation was determined as a percentage of the standard deviation of the coverage divided by the mean.

**Supplementary Figure 16:**
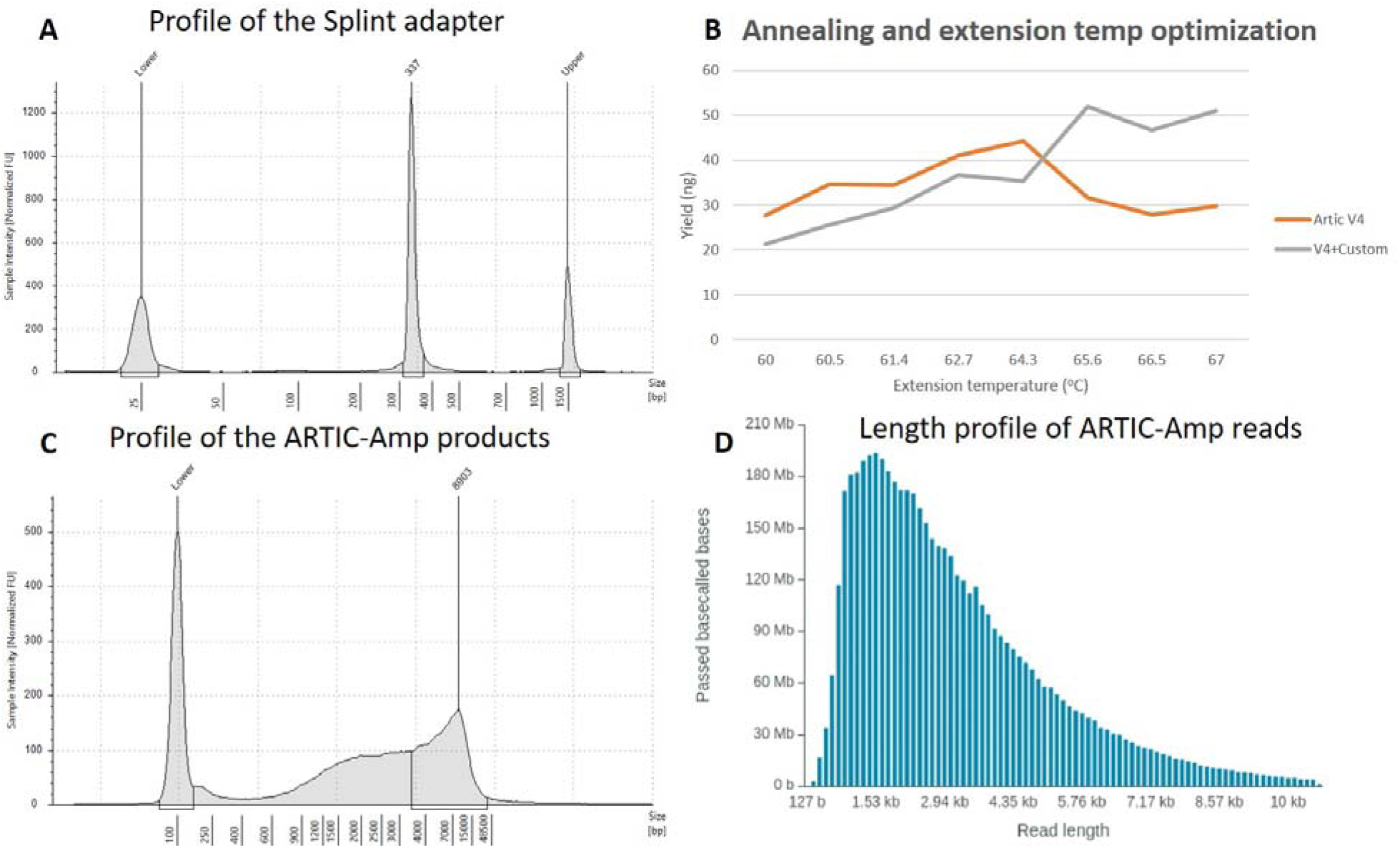
ARTIC-Amp protocol product evaluation. A) Profile of the 330 bp ‘splint’ sequence amplified. The splint is used in circularisation of ARTIC amplicons via Gibson assembly. A Tapestation-generated image is shown here (). B) Optimisation of annealing and extension temperature for the ARTIC v4.1 primers that were modified to add a 29 bp tag sequence that would enable circularisation of molecules in conjunction with the ‘splint’ via Gibson assembly. C) Profile of ARTIC-Amp final products following circularisation of amplicons and rolling circle amplification. A peak length of 8.9 kb was achieved. A Tapestation-generated image is shown here () D) Profile of reads generated from sequencing of libraries prepared through the ARTIC-Amp protocol. The image is generated by MiniKNOW (Oxford Nanopore Technologies, UK).

**Supplementary Figure 17:**
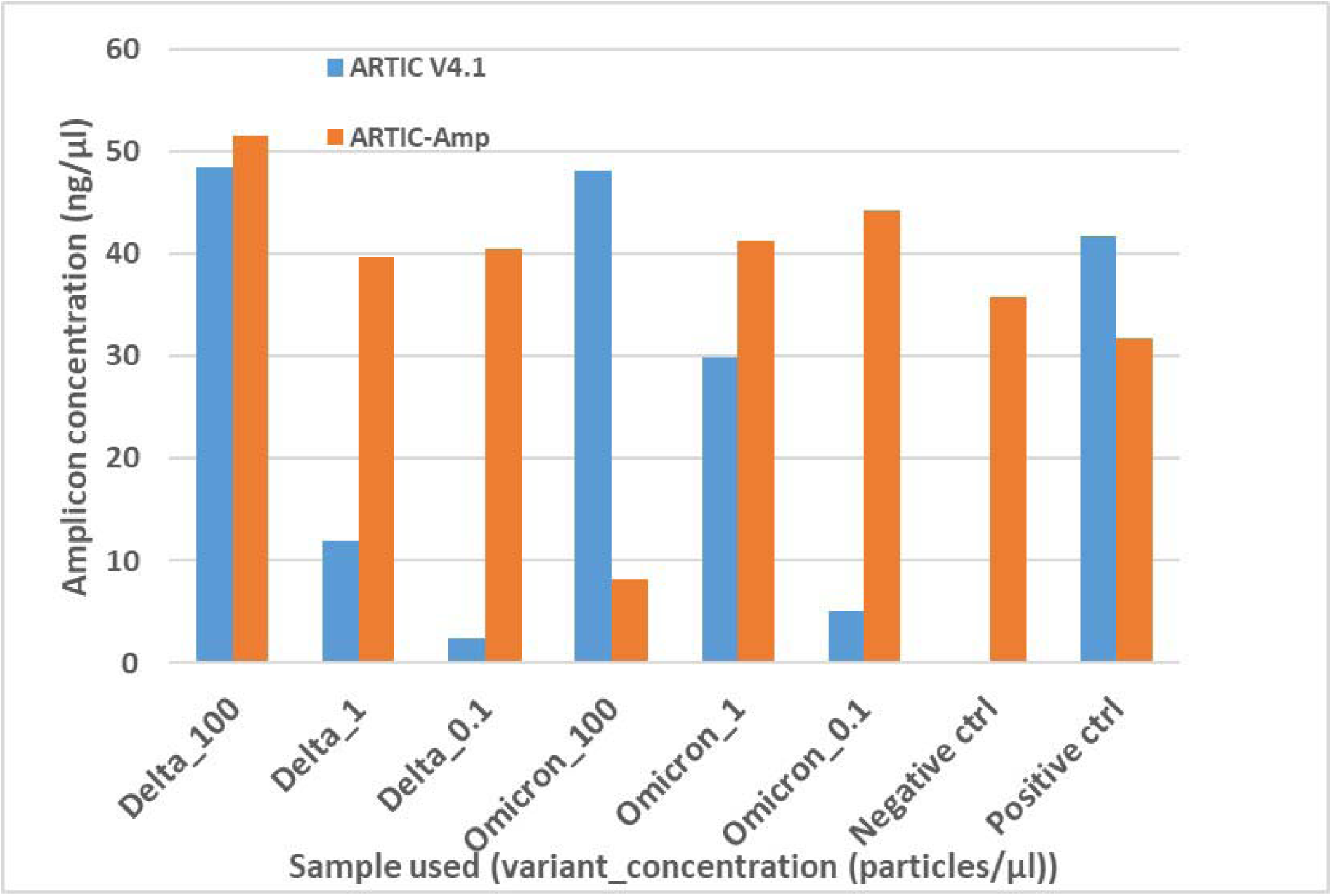
Amplicon concentration comparison between ARTIC v4.1 and ARTIC-Amp protocols. Four primer sets targeting regions in four SARS-CoV-2 genes: N, ORF7a, ORF1a, and Spike, respectively were used to prepare sequencing libraries either following the ARTIC v4.1 protocol or our inhouse protocol termed ARTIC-Amp. The ARTIC-Amp protocol takes the final products of the ARTIC v4.1 protocol and circularises them via Gibson assembly followed by isothermal rolling circle amplification as described previously (Volden et al., 2018). Two SARS-CoV-2 variant namely B.1.617.2 (Delta) and Omicron (BA.1) samples were processed at three different concentrations: 100, 1, and 0.1 particles/µL. Low viral load samples (0.1 particles/µL) were processed in triplicate and the average is shown here.

**Supplementary Figure 18:**
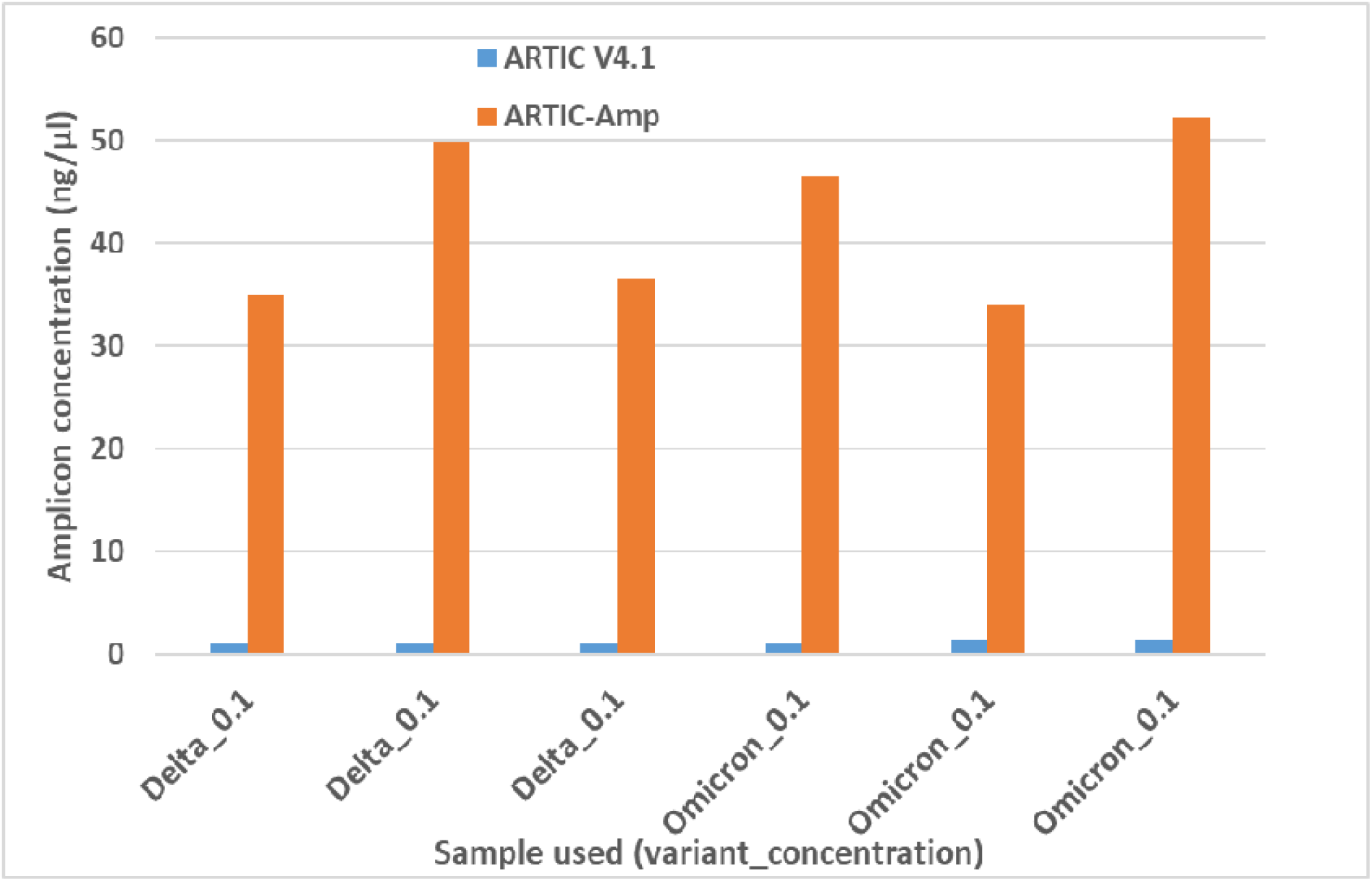
Same as Supplementary Figure 17 but showing low viral load samples (0.1 particles/µL) which were processed in triplicate.

**Supplementary Figure 19:**
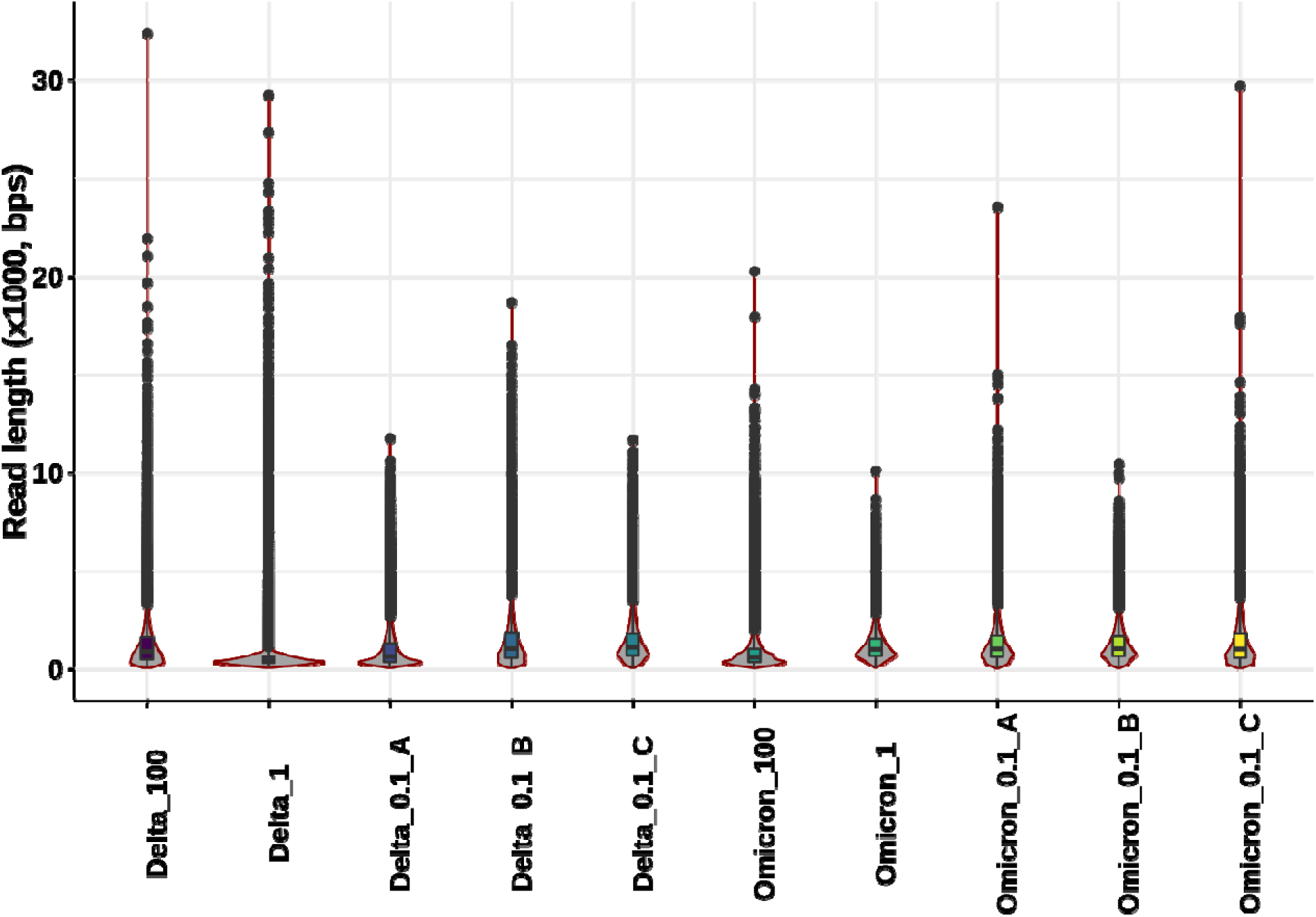
Read length distribution of ARTIC-Amp libraries. Ten samples were processed following our inhouse ARTIC-Amp protocol and sequenced on the PromethION. The reads generated were trimmed of sequencing adapters and their lengths determined. The ten samples comprised two SARS-CoV-2 cell culture variants namely: B.1.617.2 (Delta) and BA.1 (Omicron). The samples were normalised and serial diluted to three concentrations, 100, 1, and 0.1 particles/µL. Low viral load samples (0.1 particles/µL) were processed in triplicate and shown here as A, B, and C.

**Supplementary Figure 20:**
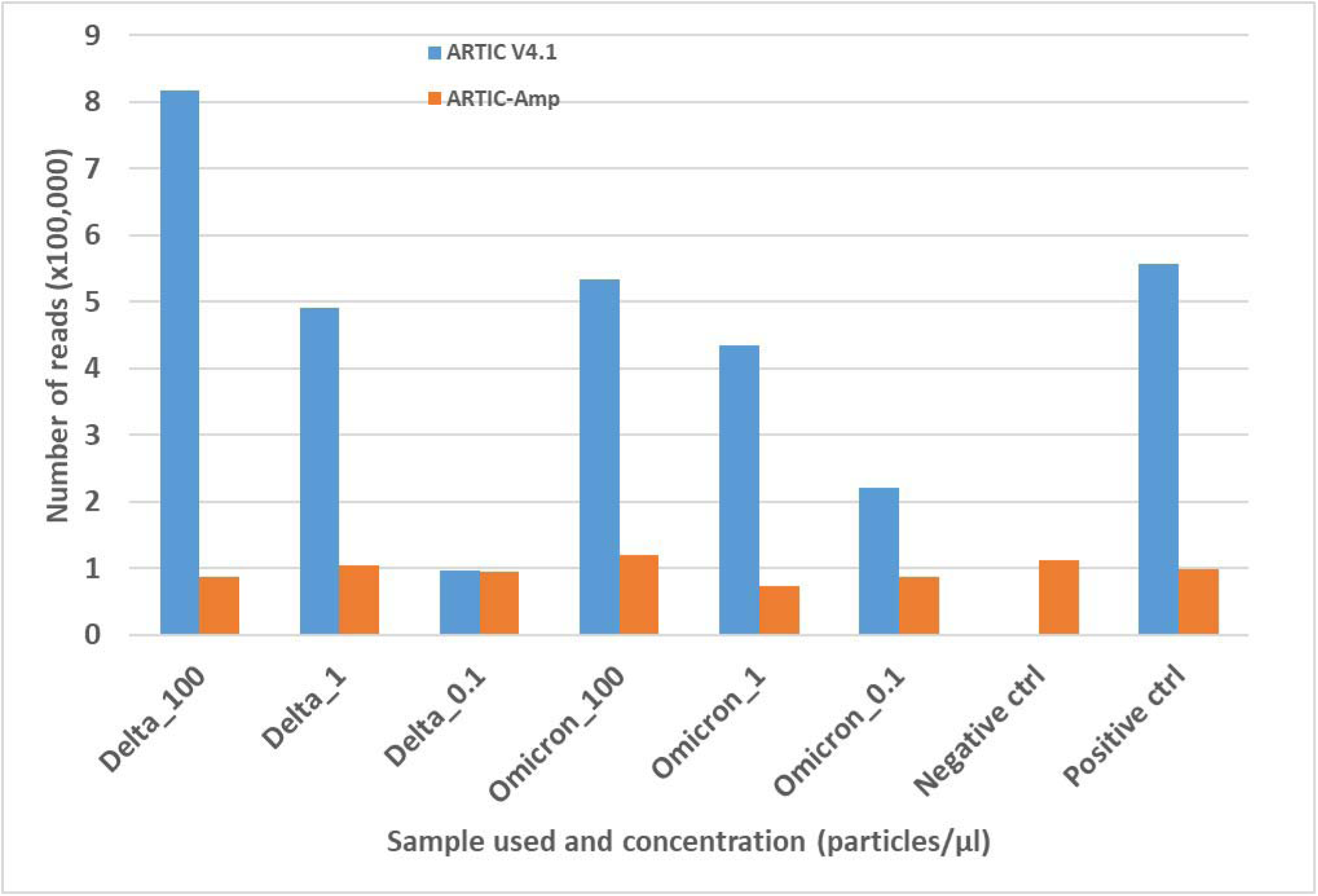
Total number of reads generated from ARTIC v4.1 and ARTIC-Amp protocols. Same details as Supplementary Figure 17 but showing total number of reads generated following sequencing on the PromethION. Low viral load samples (0.1 particles/µL) were processed in triplicate and the average is shown here.

**Supplementary Figure 21:**
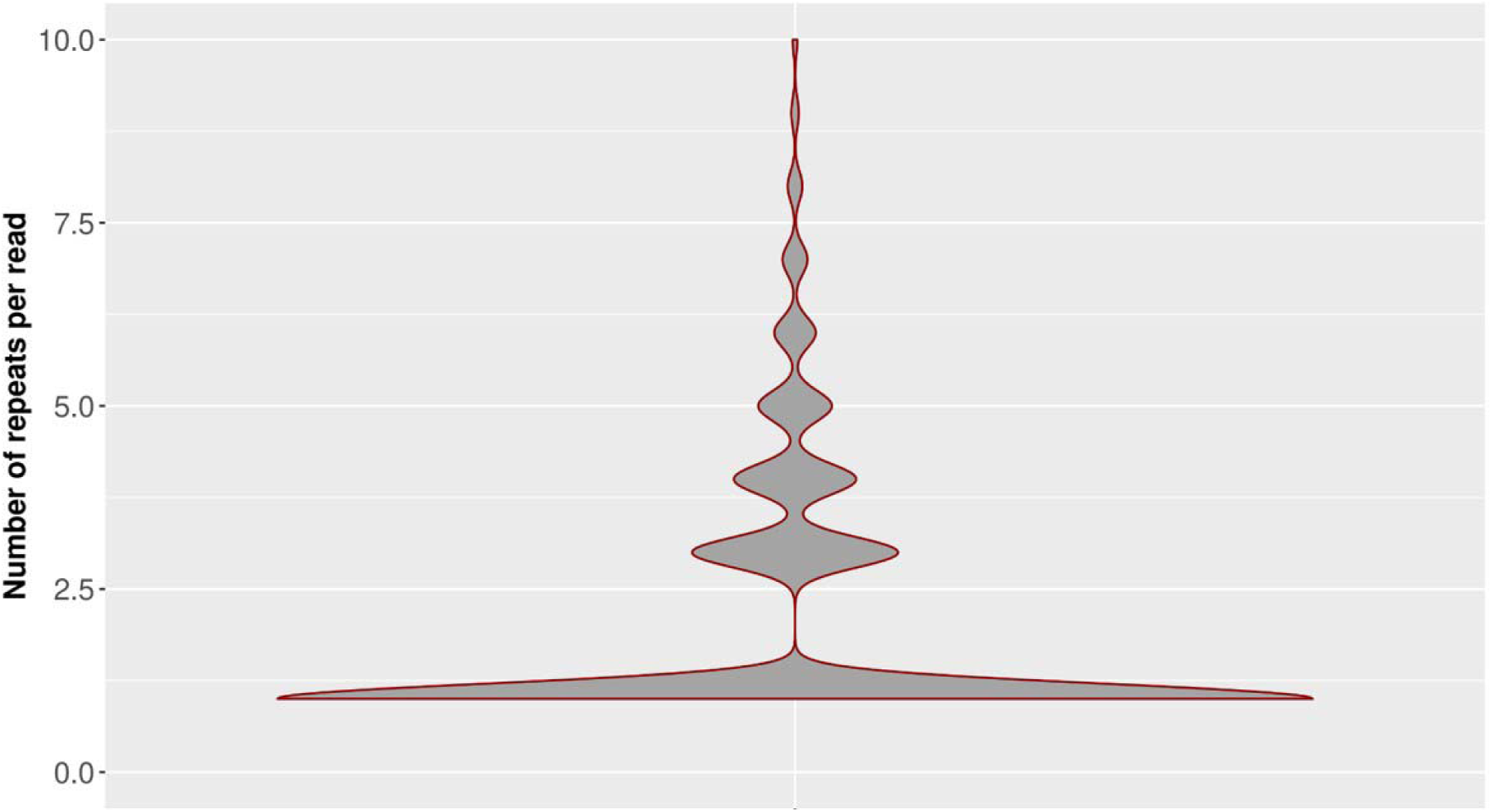
Profile of number of repeats in reads generated through ARTIC-Amp protocol. Our inhouse ARTIC-Amp protocol includes rolling circle amplification of circularised molecules which creates repeats of the circularised molecules. We used R2C2 pipeline (Volden et al., 2018) to identify and count the number of repeats in each read.

**Supplementary Figure 22:**
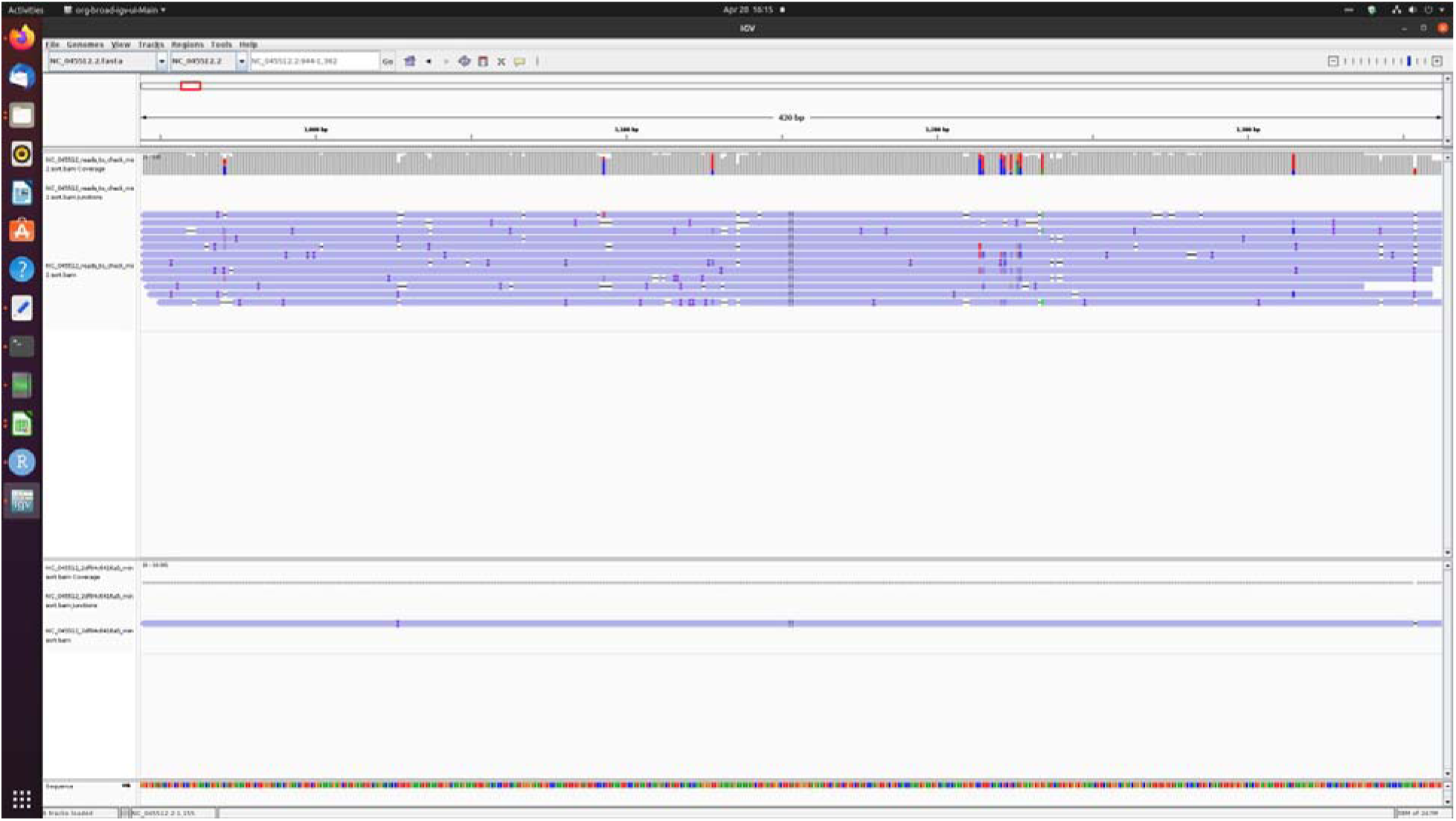
Example of consensus error correction. IGV screen shot shows in the top panel a single read that had 13 repeats and the final consensus error-corrected final read in the bottom panel.

**Supplementary Figure 23:**
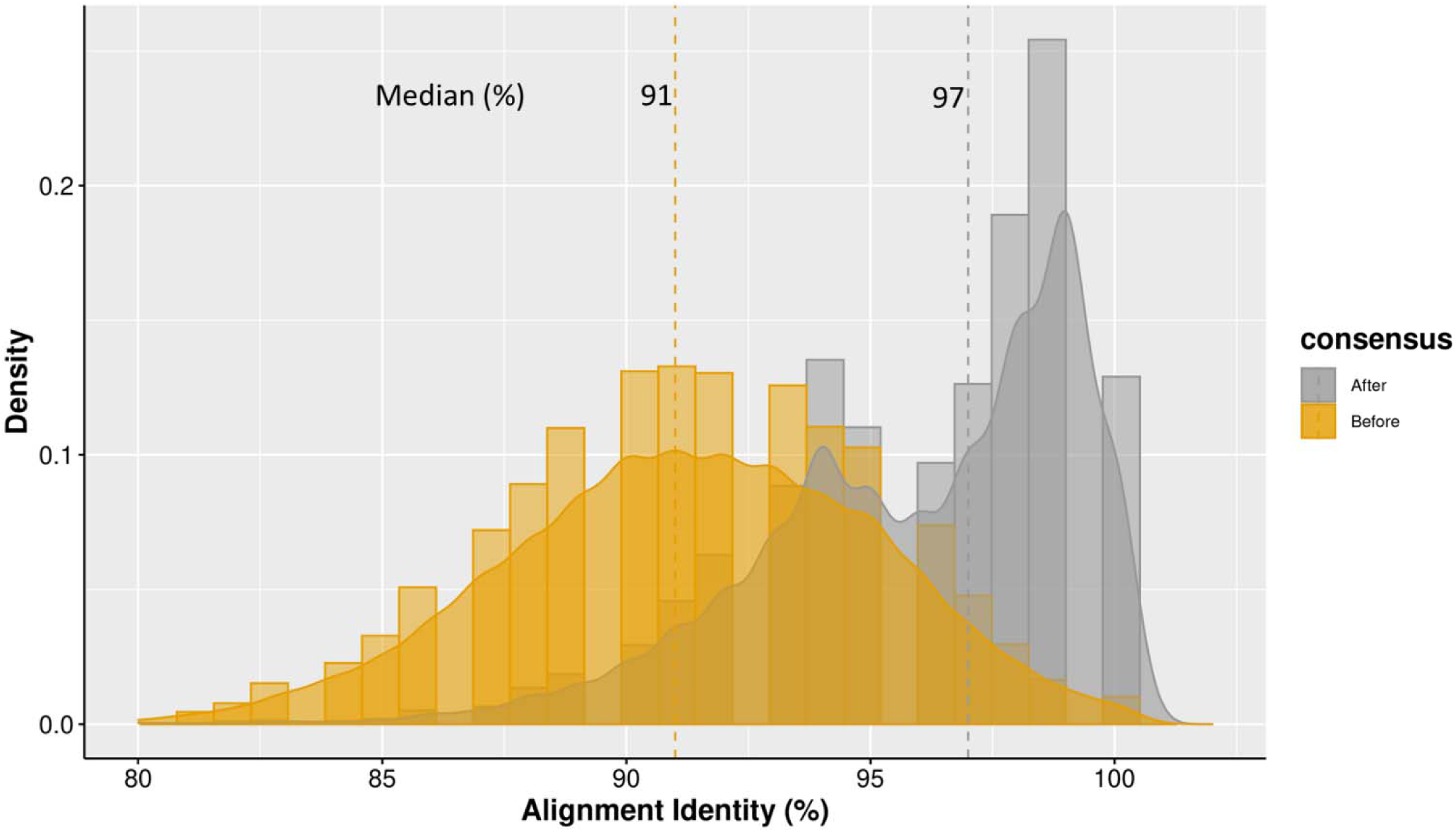
Alignment identity improvement of reads generated by ARTIC-Amp protocol. Our inhouse ARTIC-Amp protocol includes rolling circle amplification of circularised molecules which creates repeats of the circularised molecules. We used R2C2 pipeline (Volden et al., 2018) to identify repeats in each read and create a consensus error corrected final read for each molecule. The alignment identity of the reads before and after consensus error correction was determined and is shown here.

**Supplementary Figure 24:**
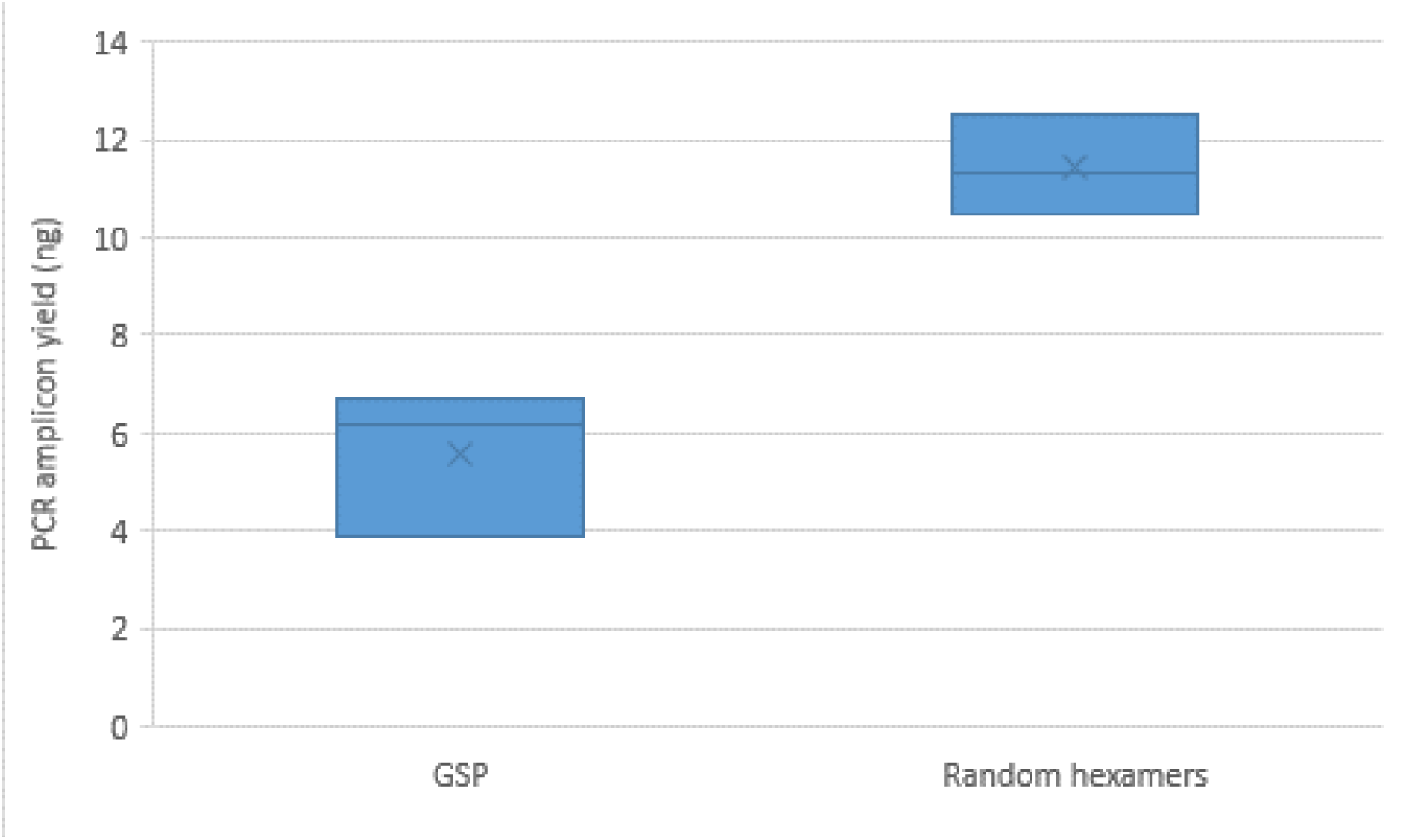
PCR amplicon yield comparison. We compared the yield from PCR amplification of cDNA either prepared using gene-specific primers (GSP) or random hexamers during reverse transcription while performing Entebbe protocol. The Entebbe protocol uses GSP while ARTIC protocol uses random hexamers.

**Supplementary Figure 25:**
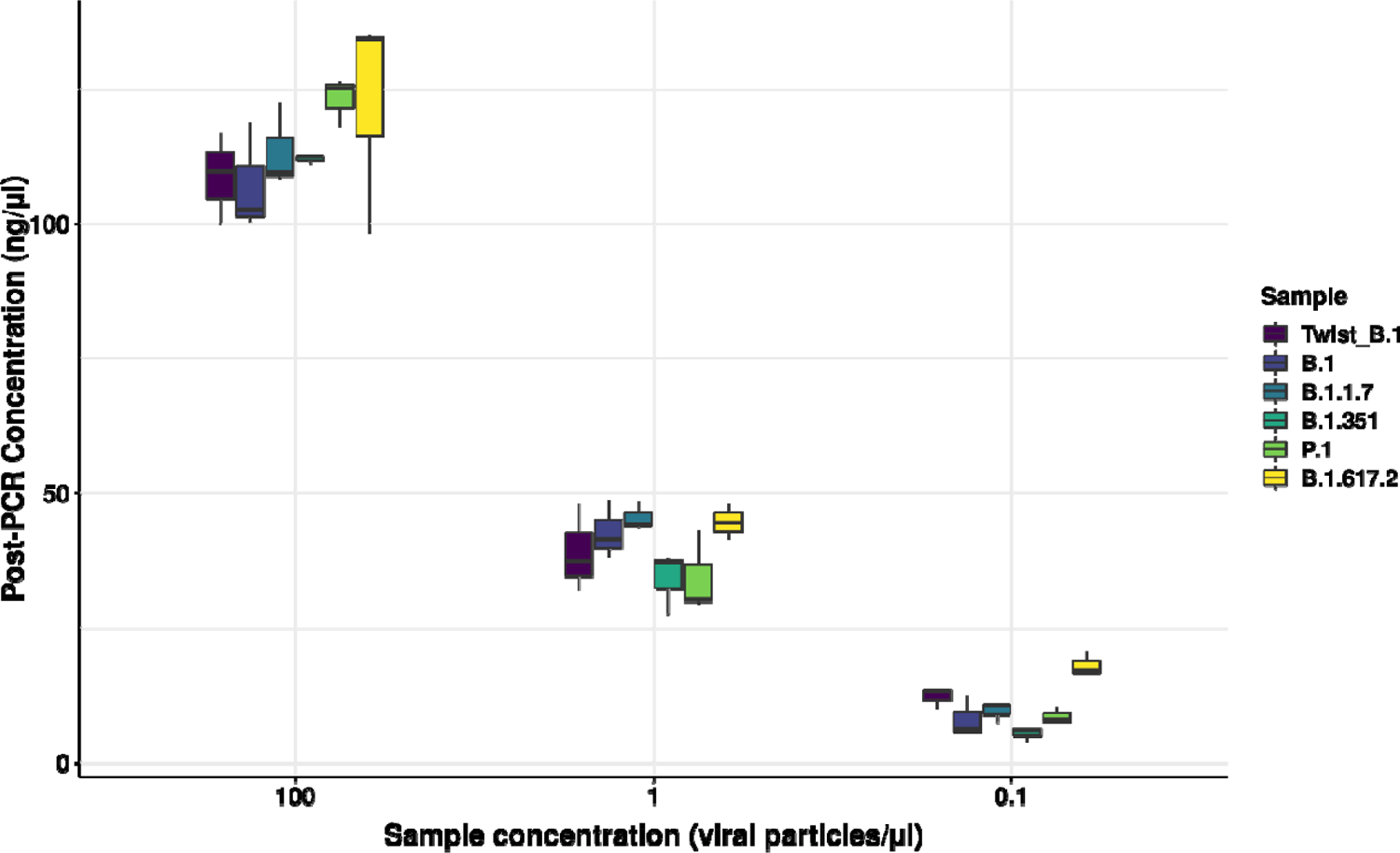
Comparing the concentration of Twist synthetic control and cell culture variants normalised by ddPCR. The Twist Synthetic SARS-CoV-2 RNA Control 1 (here shown as Twist_B.1) whose commercial concentration is 1,000,000 particles per microliter was log serial diluted to obtain samples at 1×10^5^, 1×10^3^, and 1×10^2^ particles per millilitre. We processed these samples in triplicate following the ARTIC v4 protocol and determined the post-PCR amplicon concentrations which are shown here. Cell culture wildtype and variant SARS-CoV-2 virus were purified from cell culture supernatant and their concentrations determined by digital droplet PCR (ddPCR). The cell culture samples were normalised to 1000 particles/µL and then log serial-diluted to 100, 1, and 0.1 particles per microliter. The samples were processed following the ARTIC v4 protocol and the post-PCR amplicon concentrations determined which are shown here.

