## Supplementary materials for "A Benchmark of methods for SARS-CoV-2 whole genome sequencing and development of a more sensitive method"

**Cell culture of SARS-CoV-2 variants**

**Virus production**

1. **Day -1 : (BSL2)**
   a. Seed a 175 cm^2^ flask with 10x10^6^ VERO cells/20 ml M-199 1X + 10% FBS.
   b. Incubate at 37°C, 5% CO_2_ for 24h.
2. **Day 0 (BSL3)**
   a. VERO cells should be at 80-85% confluence.
   b. Remove media from the flask.
   c. Add 15 ml of infection media + 15 ml of SARS-CoV-2 virus (approx. MOI of 0.1).
   d. Incubate at 37°C, 5% CO_2_ for 4-5 days.
   e. Infection media is M-199 1X containing 10 mM Hepes, 1 mM sodium pyruvate, 1.25 g glucose, 2.75 mg plasmocin and 2% of heat-inactivated Fetal Bovine Serum (FBS).
3. **Day 4-5 (BSL3)**
   a. Cells should be round and practically all detached from the bottom of the flask.
   b. Transfer cells and media in 50 ml conical tubes.
   c. Spin down at 500 x g for 5 minutes at 4°C.
   d. Aliquote supernatant in 2 ml tubes.
   e. Store at -80°C.

**Viral RNA purification using the Bead mill tissue RNA purification kit (26-010B, OMNI International, GA, USA).**

1. Mix 300 µL of aliquoted SARS-CoV-2 supernatant with ceramic beads, 300 µL of RLB buffer/2-mercaptoethanol and 10 µL of antifoam reagent in a special 2 ml tube.
   a) 20 µL of 2-mercaptoethanol is added per 1 ml of RLB buffer.
2. Homogenize in a Bead Mill Homoenizer (Omni International): 4 m/s for 30 seconds.
3. Centrifuge tubes at 10000 x g, 5 minutes at 4°C.
4. Transfer the cleared supernatant to a 1.5 ml microcentrifuge tube.
5. Add 1 volume of 70% ethanol.
6. Vortex for 10 seconds.
7. Insert an Omni RNA Mini Column into a 2 ml Collection Tube.
8. Transfer 700 µL of the sample to the Omni RNA Mini Column.
9. Spin at 10000 x g for 1 minute at room temperature.
10. Discard the flow-through and re-use the collection tube.
11. Repeat steps 8-9-10 until the entire sample has been transferred.
12. Add 500 µL of RW1 buffer into the column.
13. Centrifuge at 10000 x g for 30 seconds (4°C). Discard the flow-through and collection tube.
14. Add 500 µL of RW2 buffer into the column.
15. Centrifuge at 10000 x g for 1 minute (4°C). Discard the flow-through and re-use the collection tube.
16. Add another 500 µL of RW2 buffer into the column.
17. Centrifuge at 10000 x g for 1 minute (4°C). Discard the flow-through and re-use the collection tube.
18. Centrifuge at 10000 x g for 2 minutes (4°C) to completely dry the column.
19. Place the Omni RNA Mini Column onto a 1.5 ml microcentrifuge tube and add 50 µL of DEPC-treated water.

Centrifuge at 10000 x g for 2 minutes (4°C) to elute RNA.

**Lambda phage splint**

**DNA Splint Amplification from Lambda DNA**

1. Make 10 μM of Lambda_Splint_F and Lambda_Splint_R
2. Label 3 different as W, X, and Y
3. Prepare the following mastermix

| **No.** | **Component** | **Volume/sample (µL)** | **V4/Cus_Mix (vol, µL)**  **4 samples** |
| --- | --- | --- | --- |
| 1. | Q5 Hot Start High-Fidelity 2X Mix | 12.5 | 50 |
| 2. | Lambda_Splint_F (10 μM) | 0.5 | 2 |
|  | Lambda_Splint_R (10 μM) | 0.5 | 2 |
| 3. | Nuclease-free water | 6.5 | 26 |
|  | **Total Volume** | **20** |  |

4. Add 20 µL to each of the tubes W, X, Y, and Z (Z is negative ctrl)
5. Add 5 µL of Lambda DNA to the corresponding tubes and water in the negative control.
6. Amplify the Splint as follows

| **Step** | **Cycles** | **Temp (°C)** | **Time (minutes)** |
| --- | --- | --- | --- |
| Heat Activation | 1 | 95 | 3 |
| Denature | 25 | 98 | 20 sec |
| Annealing |  | 67 | 15 sec |
| Extension |  | 72 | 30 sec |
| Hold |  | 4 | ∞ |

7. Pool all Lambda Splint amplicons and do 1X AMPure clean up and elute the DNA in 15 ul water.
8. Perform quality control (QC). Quantitate and profile with a Tapestation 4200 analyzer with the D5000 assay kit as well as the Qubit fluorimeter.

Qubit quantificaton

| **Code** | **I.D** | **vol** | **Dil** | **µL Qbt** | **Conc’n (ng/µL)** | **Yield (ng)** |
| --- | --- | --- | --- | --- | --- | --- |
| WXY | Lambda 1 | 15 | 10 | 1 | 23.2*10 = 232 | 3480 |

Tapestation D5000 HS assay profile

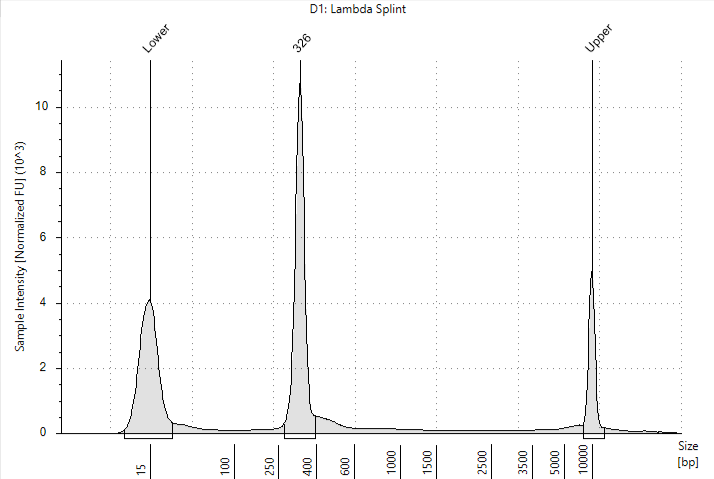

**Custom protocol**

**R2C2 circularization**

1. Make the following R2C2 sample preparation mix

| No. | **Component** | **Volume/sample (µL)** | **x samples** |
| --- | --- | --- | --- |
| 1. | Water | 12-(x+y) |  |
| 2. | cDNA (<=200 ng) | X |  |
| 3. | Splint DNA (<=600 ng) | y |  |
| 4. | 2x NEBuilder HiFi DNA Assembly Master Mix | 6 |  |
|  | **Total Volume** | **12** |  |

2. Incubate at 55 °C for 60 minutes
3. Remove non-circularized cDNA as follows

| No. | **Component** | **Volume/sample (µL)** | **x samples** |
| --- | --- | --- | --- |
| 1. | Water | 4 |  |
| 2. | Lambda Exonuclease Reaction Buffer (10X) | 2 |  |
| 3. | cDNA-Splint rxn | 12 |  |
| 4. | Exo_III:Lambda_Exo* | 1 |  |
| 5. | Exonuclease I | 1 |  |
|  | **Total Volume** | **20** |  |

*Exonuclease III and Lambda Exonuclease are prepared in 1:10 ratio

4. Incubate at 37°C for 30 minutes and heat inactivate at 75°C for 10 min.
5. Extract circularized DNA using 1.2X SPRI beads (24 ul) purification to keep as many of the fragments ~500 bp and elute in 10 μL of ultrapure water.

**Rolling circle amplification**

6. Perform Rolling circle amplification as follows

| No. | **Component** | **Volume/sample (µL)** | **x samples** |
| --- | --- | --- | --- |
| 1. | Water | 29 |  |
| 2. | 10× Phi29 Buffer | 5 |  |
| 3. | dNTPs 10 µM (each) | 2.5 |  |
| 4. | Exonuclease resistant random hexamers (10 µM) | 2.5 |  |
| 5. | Phi29 | 1 |  |
| 6. | Circularized cDNA | 10 |  |
|  | **Total Volume** | **50** |  |

7. Incubate reaction at 30 °C overnight. (Started at 16:23 on 30^th^.03.22, Stopped at 10:00 on 31^st^.03.22, total of 17 hours 37 minutes)
8. Adjust volume to 300 μL with ultrapure water.
9. Extract DNA using SPRI beads with a size cut-off to eliminate DNA <2,000 bp (0.5 beads:1 sample).
   a. Use the following elution buffer to elute and de-branch the DNA

| No. | **Component** | **Volume/sample (µL)** | **x samples** |
| --- | --- | --- | --- |
| 1. | NEB buffer 2 | 10 |  |
| 2. | Water | 90 |  |
| 3. | T7 Endonuclease | 5 |  |

b. Incubate beads for two hrs on a thermal shaker at 37 °C under constant agitation. I incubated in 1.5 ml tubes, spinning at 1000 rpm on … in room 5300.
c. Recover the supernatant by placing the tubes containing the beads on magnets

10. Perform final SPRI beads purification with a size cut-off to eliminate DNA <2,000 bp (0.5 beads:1 sample) and elute in 15 μL of ultrapure water
11. Perform quality control (QC). Quantitate (A Tapestation 4200 analyzer with the D5000 assay kit as well as the Qubit fluorimeter were used.)
12. Adjust the amount of DNA in the tube to be 100 ng/μL and give samples to Production team for Nanopore library preparation and sequencing.

**Cost of R2C2**

|  | **R2C2 circularization** | **cost (USD)/reaction** | **Catalog number** |
| --- | --- | --- | --- |
| 1 | 2x NEBuilder HiFi DNA Assembly Master Mix | 6.21 | E2621S |
|  | **Remove non-circularized cDNA** |  |  |
| 2 | Lambda Exonuclease | 5.20 | M0262S |
| 3 | Exonuclease III | 1.86 | M0206S |
| 4 | Exonuclease I | 0.69 | M0293S |
|  | **Rolling circle amplification** |  |  |
| 5 | Exonuclease resistant random hexamers (10 µM) | 0.05 | SO181 |
| 6 | Phi29 DNA polymerase | 3.44 | M0269S |
|  | **De-branching** |  |  |
| 7 | T7 Endonuclease | 3.47 | M0302S |
|  | **Total cost per reaction** | **20.92** |  |
